# Targeting phenotypic plasticity prevents metastasis and the development of chemotherapy-resistant disease

**DOI:** 10.1101/2022.03.21.22269988

**Authors:** Beatriz P San Juan, Soroor Hediyeh-Zadeh, Laura Rangel, Heloisa H Milioli, Vanina Rodriguez, Abigail Bunkum, Felix V Kohane, Carley A Purcell, Dharmesh D Bhuva, Anie Kurumlian, Lesley Castillo, Elgene Lim, Anthony J Gill, Vinod Ganju, Rachel Dear, Sandra O’Toole, A. Cristina Vargas, Theresa E Hickey, Leonard D Goldstein, John G Lock, Melissa J Davis, Christine L Chaffer

**Author notes:** Correspondence: Christine Chaffer, Melissa Davis, Beatriz Pérez San Juan.

## Abstract

Cancer cells invoke phenotypic plasticity programs to drive disease progression and evade chemotherapeutic insults, yet until now there have been no validated clinical therapies targeting this process. Here, we identify a phenotypic plasticity signature associated with poor survival in basal/triple-negative breast cancer, in which androgen signalling is prominent. We establish that anti-androgen therapies block cancer stem cell function and prevent chemotherapy-induced emergence of new cancer stem cells. In particular, the anti-androgen agent seviteronel synergizes with chemotherapy to improve chemotherapeutic inhibition of primary and metastatic tumour growth and prevent the emergence of chemotherapy-resistant disease. We validate cytoplasmic AR expression as a clinical phenotypic plasticity biomarker that predicts poor survival and poor response to chemotherapy, and positive response to seviteronel plus chemotherapy. This new targeted combination therapy validates modulating phenotypic plasticity as an effective strategy to prevent and treat chemotherapy-resistant cancers with transformative clinical potential.

**STATEMENT OF SIGNIFICANCE:** There are currently no curative therapies for patients with chemotherapy-resistant cancer. We demonstrate that modulating phenotypic plasticity prevents the emergence of chemotherapy-resistant disease in triple-negative breast cancer. This represents the first known validated clinical therapy leveraging phenotypic plasticity. Moreover, we identify a highly effective anti-androgen drug and a biomarker to select and treat patients best-suited to this new therapy. A clinical trial is underway (NCT04947189).

**SUMMARY SENTENCE:** Blocking phenotypic plasticity is an effective targeted therapeutic strategy to treat cance

## INTRODUCTION

For decades, studies have demonstrated the importance of phenotypic plasticity in driving cancer progression to the point that it is now recognized as a hallmark of cancer ^1^. Nonetheless, there are currently no clinically approved therapies that exploit modulation of cell plasticity to improve survival outcomes for cancer patients.

Phenotypic plasticity is a molecular process that enables cancer cells to transition between distinct cell states in a rapid and dynamic manner. The phenotypic flexibility imparted by this process enables cancer cells to adapt and thrive in otherwise arduous conditions, including foreign microenvironments encountered at distant sites of metastasis or to evade a mounting immune response ^2–6^. Of particular interest, phenotypic plasticity enables cancer cells to evade chemotherapeutic insults ^7, 8^. These data suggest that targeting phenotypic plasticity may be an effective strategy to prevent and treat metastatic and chemotherapy-resistant disease ^9–12^.

Many insights into the molecular drivers, and thus, potential therapeutic targets, of phenotypic plasticity in cancer have been garnered through studies of the epithelial-to-mesenchymal transition (EMT). In cancer, the EMT enables poorly aggressive epithelial cancer cells to transition into the highly aggressive cancer stem cell state that is associated with increased metastatic and chemotherapy-resistant capabilities ^13–17^. The EMT program is underpinned by a core transcriptional circuitry involving SNA1/2, ZEB1/2 and TWIST1/2 that may be upregulated in response to contextual signals arising in the tumour microenvironment ^15^. We have also learned that epigenetic regulation plays an important role in cell plasticity^18^. For example, some epithelial cancer cells are predisposed to undergoing the EMT by maintaining the promoter of core EMT transcription factors in a bivalent chromatin configuration to facilitate rapid gene activation ^14, 18^.

The co-option of plasticity programs in cancer has important therapeutic implications. We now appreciate that chemotherapy can activate the EMT program^19^. This means that chemotherapy itself facilitates the development of chemotherapy-resistant disease ^16, 17, 19^. In those cases, effective therapeutic strategies for patients will require a two-pronged approach that combines standard-of-care chemotherapy with a targeted therapy to inhibit plasticity programs.

A critical limitation to the clinical translation of therapies targeting plasticity is that it is not yet possible to clinically identify or quantitate a tumour’s propensity to invoke a cell plasticity program. Accordingly, identifying tumor biomarkers of cell plasticity, and targeted therapies to match to those patients, remains an unmet need.

It may, however, be possible to discern the clinical manifestation of plasticity programs. For example, response to chemotherapy in patients with triple-negative breast cancer is broadly dichotomous: whereas two thirds of patients obtain a pathological complete response associated with good prognosis and overall survival, one third of patients respond poorly and inevitably develop recurrent and chemotherapy-resistant disease ^20, 21^. Overall survival is typically less than 18 months for those patients ^22.^ These data highlight two current clinical challenges: 1) the inability to identify patients at diagnosis who will not obtain a pathological complete response to primary chemotherapy, and, 2) the lack of effective treatment options for those patients when standard-of-care treatments fail.

We hypothesised that strategies to prevent phenotypic plasticity will prove transformative to clinical management and survival outcomes for cancer patients. We previously demonstrated that dynamic regulation of the EMT transcription factor ZEB1 is essential for cancer cell plasticity programs in triple-negative breast cancer, whereby ZEB1 activation drives cancer cells into a cancer stem cell state, and ZEB1 inactivation is required for re-differentiation and efficient growth of metastatic lesions ^14^. With no way yet to therapeutically target ZEB1, translation of a viable therapeutic strategy to stop phenotypic plasticity has not been possible. Accordingly, we set out to identify and validate therapeutic strategies targeting phenotypic plasticity to prevent and treat chemotherapy-resistant disease. Herein, we show that androgen signalling is a driver of phenotypic plasticity in triple-negative breast cancer. Furthermore, we validate the novel application of anti-androgen therapies as clinical inhibitors of phenotypic plasticity induced by chemotherapy. Thus, the combination of anti-androgen therapy in combination with standard-of-care chemotherapy significantly inhibits primary, metastatic and chemotherapy-resistant disease. Moreover, we identify that cytoplasmic expression of the androgen receptor is a predicative and prognostic biomarker for poor response to chemotherapy, and can thus be used to identify patients best-suited to this novel targeted combination therapy.

## RESULTS

### Identifying therapeutically targetable drivers of phenotypic plasticity in triple-negative breast cancer

We sought to construct the transcriptomic network that governs entrance into, and maintenance of, the aggressive and chemotherapy-resistant CD44^Hi^ cancer stem cell state. We first isolated CD44^Lo^ and CD44^Hi^ populations from TNBC cell lines (HMLER and HCC38) through consecutive rounds of FACS enrichment (Fig. 1A-B; Supplementary Fig. S1A). Echoing previous work ^14, 23, 24^ CD44^Hi^ cells reside in a partial-EMT state (high ZEB1/low E-cadherin) and displayed greater tumorsphere-forming capacity compared to their CD44^Lo^ counterparts (Supplementary Fig S1B-C). For comparative purposes, we used two luminal, estrogen receptor positive, breast cancer cell lines (MCF7 and ZR-751) as examples of CD44^Lo^ cells that do not readily switch states ^14^. The latter cell lines do not endogenously harbour a CD44^Hi^ cell population.

**Figure 1:**
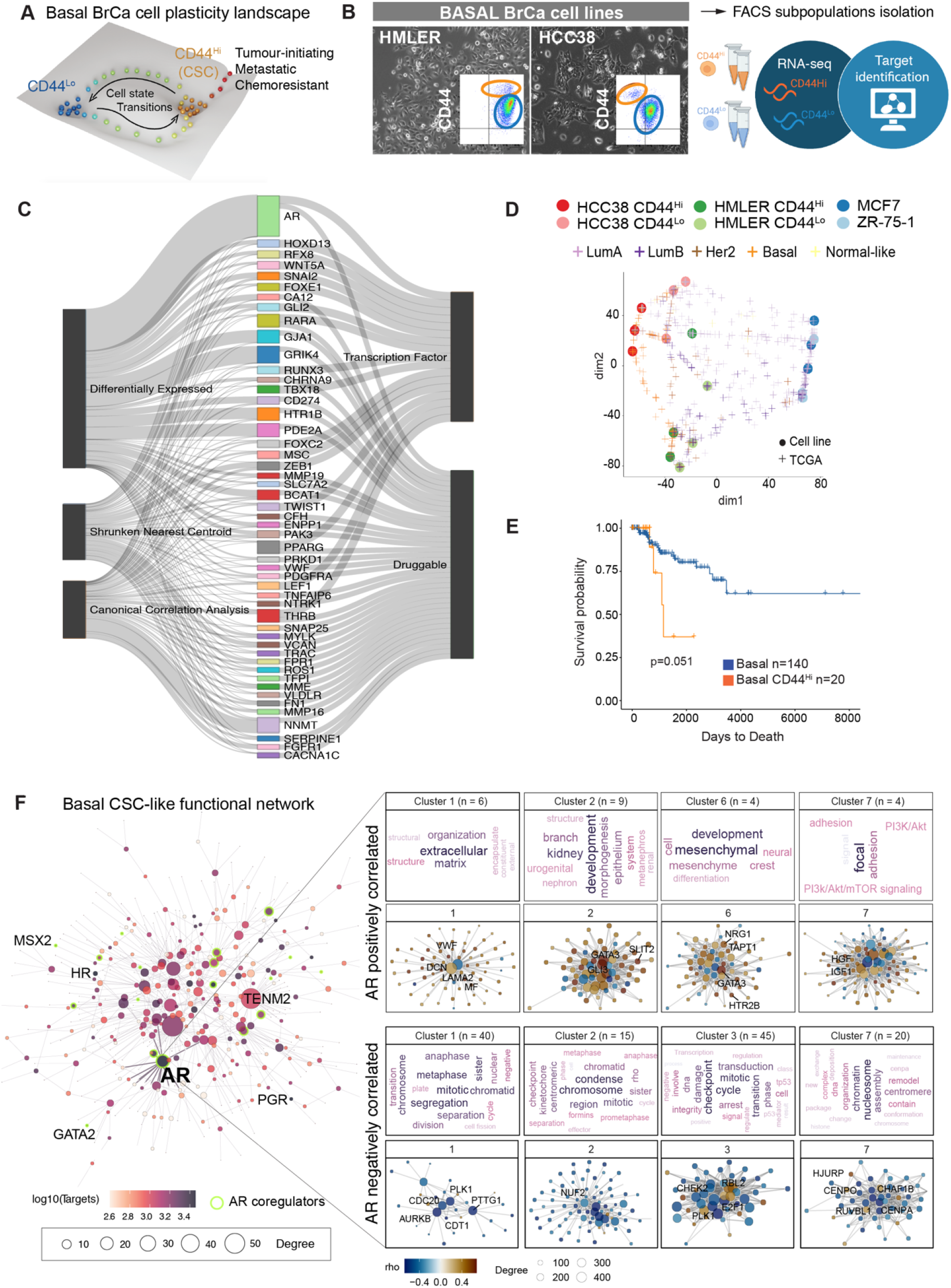
Identifying therapeutically targetable drivers of phenotypic plasticity in triple-negative breast cancer. **A**, Schematic illustrating cancer cell heterogeneity and cancer cell state transitions across the basal breast cancer cell plasticity landscape. **B**, Photomicrographs showing morphology and CD44 FACS profile (insert) of basal cell lines used for FACS isolation of subpopulations, RNA-seq and identification of cell plasticity targets. **C**, Sankey plot of discriminative and correlative analyses performed on transcriptome (RNA-seq) of purified triple-negative breast cancer CD44^Lo^ and CD44^Hi^ subpopulations. **D**, Optimal Transport (OT) map cell line representing correspondence between cell line transcriptomes and TCGA breast cancer transcriptomes. **E**, Kaplan-Meier survival outcomes of CD44^Hi^-like TCGA samples compared to all other basal breast cancer samples determined by OT correspondence analysis (D). **F**, A differential protein regulatory network of signalling pathways enriched in CD44^Hi^-like TCGA breast cancer samples.

RNA-seq was performed on the purified cell sub-populations, followed by differential expression, canonical correlation and nearest shrunken centroid analyses (Fig. 1C; Supplementary Fig. S1D). To facilitate rapid clinical implementation of selected targets, we sought to identify differentially expressed transcription factors for which there are existing drug compounds. These analyses identified AR, GRIK4, NNMT, GJA1, RARA, THRB PPARG, PDE2A, HTR1B and ZEB1 among the top mediators of the basal CD44^Hi^ state, and thus as potential drivers of CD44^Lo^ to CD44^Hi^ cancer cell plasticity (Fig. 1C).

To determine the clinical importance of those targets, we tested for a relationship between the basal cell line CD44^Hi^ signatures with clinical breast cancer specimens. We used a data projection method, Optimal Transport (OT), to map transcriptomes of basal and luminal cell line subpopulations to patient samples in the TCGA clinical breast cancer cohort ^25^. This method identifies dimensions of similarity in the two datasets and maps both data types to a common frame of reference. We used a nearest neighbour approach to identify basal CD44^Hi^-like, basal CD44^Lo^-like, or luminal CD44^Lo^-like TCGA samples (Fig. 1D). Most basal samples in TCGA map to basal cell lines, particularly to the CD44^Hi^ subpopulation. Most Luminal TCGA samples mapped to the luminal CD44^Lo^ cell subpopulations. Kaplan-Meier analysis demonstrated that TCGA samples mapped closest to CD44^Hi^ cells (n=20) had reduced overall survival compared to all other basal samples (n=140, p=0.051; Fig. 1E). These data demonstrate that the transcriptome of aggressive basal CD44^Hi^ cells identifies a clinical subset of basal breast cancers with poor outcome.

To determine dominant signalling pathways in the TCGA CD44^Hi^-like tumors we derived a protein differential regulatory network using DCANR ^26^ comparing TCGA CD44^Hi^-like to TCGA CD44^Lo^-like samples (Fig. 1F; Supplementary Fig. S1E). Transcription factors and cofactors were ranked by the number of targets regulated in either the CD44^Hi^ or CD44^Lo^ networks. Of note, an AR signalling network ranked the highest in TCGA CD44^Hi^ –like samples (Fig. 1F).

Pathways and processes identified as positively correlated with AR include mesenchymal development, organisation of the extracellular matrix and regulation of focal adhesions with evidence for signalling through ZEB1, GATA2 and the nuclear receptor NR2F1 (Fig. 1G; Supplementary Fig. S1F). Pathways and processes identified as negatively correlated with AR in CD44Hi-like samples include chromatin structure and RNA processing (Supplementary Fig. S1G). In contrast, CD44^Lo^-likeTCGA samples were dominated by a TCF transcription factor differential network (Supplementary Fig. S1E).

Together, these data identify an AR-regulatory network in clinical TNBC data that is associated with poor survival.

The AR is a steroid hormone receptor that drives context-specific transcriptional programs that regulate cell cycle, growth and metabolic pathways ^27^. Aberrant AR signalling is an unquestionable oncogenic driver of prostate cancer, but the role of AR in different breast cancer subtypes and disease stages has been highly controversial ^28^, leading to attempted clinical implementation of both AR agonists and AR antagonists in clinical trials with limited clinical impact ^29–31^. Importantly, its role as a cancer cell plasticity driver is not defined in TNBC, hence we decided to further analyse its potential as novel therapeutic modulator of cancer cell plasticity.

### Inhibiting AR signalling blocks CD44^Hi^cancer stem cell function

Immunofluorescent analysis and quantification showed low AR expression in CD44^Lo^ cells, and significant up-regulation of AR in CD44^Hi^ cells (Fig. 2A-B). High AR expression was confirmed in two additional cell lines that have a CD44^Hi^ profile (MDA-MB-231 and SUM159PT; Supplementary Fig. S2A-B). Of note, AR and ZEB1 expression were concomitantly enriched in basal CD44^Hi^ cells (Fig S2A-B). For comparative purposes, levels of AR expressed in basal HCC38 and HMLER cells are significantly lower than AR expression in luminal breast cancer cell lines (MCF7 and ZR-75-1; Supplementary Fig. S2C).

**Figure 2:**
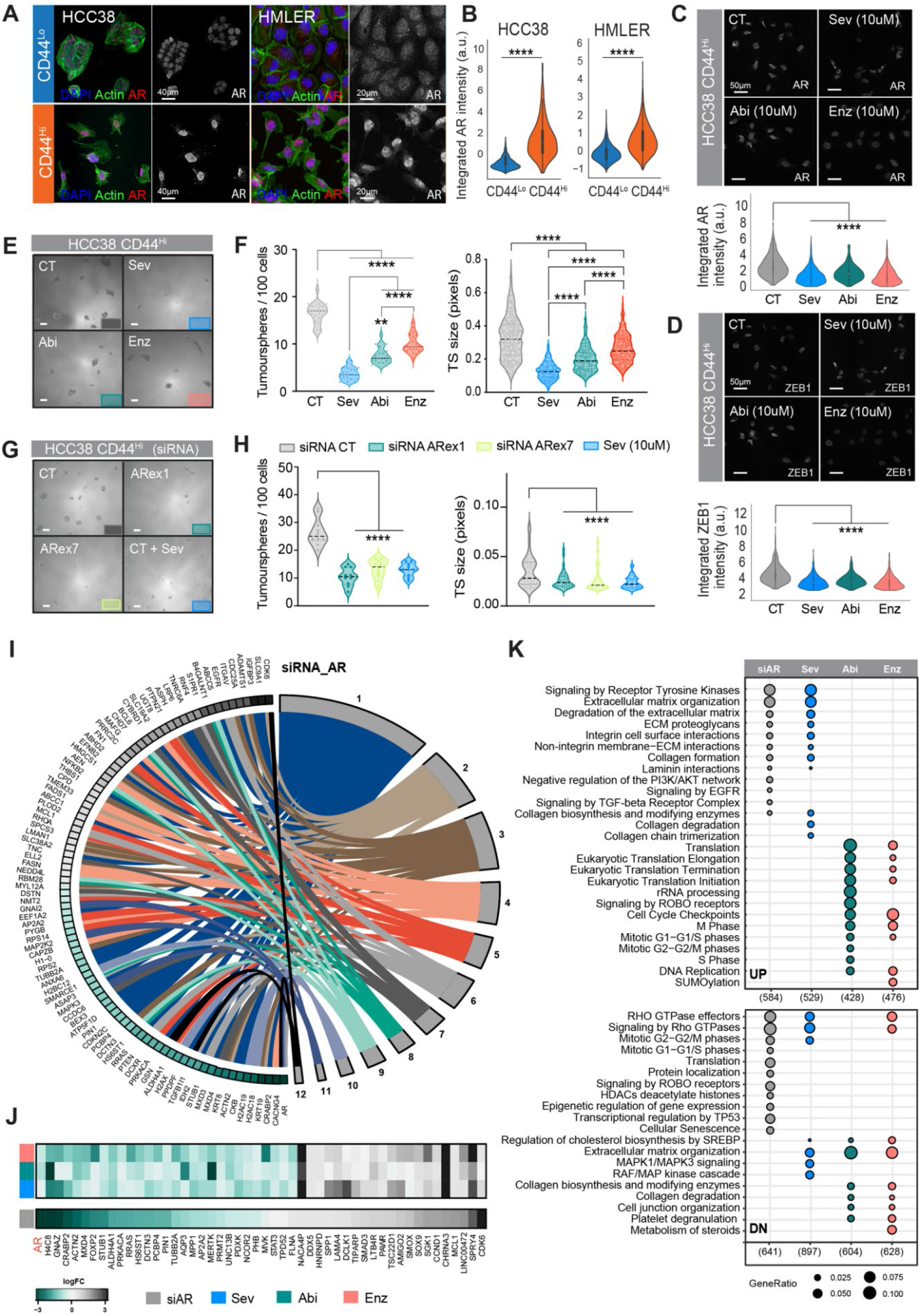
Anti-androgen agents inhibit cancer stem cell function. **A**, Immunofluorescence (IF) staining for AR (red), actin (green) and DAPI (blue) in HCC38 and HMLER CD44^Lo^ and CD44^Hi^ cell subpopulations. **B**, Quantification of AR levels from A. **C**, IF staining and quantification (violin plots) for AR in CD44^Hi^ cells following treatment with control (DMSO) or 10 μm AR inhibitors (seviteronel, abiraterone and enzalutamide). **D**, IF staining and quantification (violin plots) for ZEB1 in CD44^Hi^ cells following treatment with control (DMSO) or 10 μm AR inhibitor. **E**, Representative images of tumorsphere formation in HCC38 CD44^Hi^ cells following treatment with DMSO, Sev, Abi or Enz. Scale bars, 200 μm. **F**, Quantification of tumorsphere number and size from E. **G**, Representative images of tumorsphere formation in HCC38 CD44^Hi^ cells treated with siRNA-CT, siRNA-ARex1-ex7 or Sev + siRNA_CT. Scale bars, 200 μm. **H**, Quantification of tumorsphere number and size form G. **I**, AR TNBC regulated gene set defined by siAR compared to siCT, matched across MSigDB AR signatures (minimum of four overlapping genes. **J**, Heat map of AR-target genes regulated by siAR compared to siCT, and 10 μM Sev, Abi or Enz compared to control. **K**, Gene set enrichment analysis showing similarities and differences in up and down-regulated pathways from siAR compared to siCT or Sev, Abi or Enz compared to control.

To determine whether inhibition of AR is a potential therapeutic means of eradicating basal CD44^Hi^ cells, we investigated three drugs that antagonize AR activity: Enzalutamide, abiraterone and seviteronel. Enzalutamide is a direct AR antagonist FDA approved for treatment of prostate cancer ^32^. Abiraterone is a steroidal anti-androgen and is also FDA approved for prostate cancer ^33^, while seviteronel (VT-464/INO-464) is a new, non-steroidal anti-androgen that is not yet FDA approved for cancer treatment^34^. Abiraterone and seviteronel are both CYP17lyase inhibitors that impede production of androgenic steroids, including testosterone, a natural ligand of the AR. All three drugs reduced total AR and ZEB1 protein expression in CD44^Hi^ basal cells (Fig. 2C-D), and inhibited viability of CD44^Hi^ cells, but only at doses above their respective IC50s (Supplementary Fig. S2D).

To test whether AR antagonists could inhibit CD44^Hi^ cancer stem cell function, we used the tumorsphere assay, a surrogate *in vitro* test for *in vivo* tumour-initiating potential ^13, 35^. While all three anti-androgenic drugs significantly reduced tumorsphere formation and size, seviteronel was consistently more effective than either abiraterone or enzalutamide (Fig. 2E-F; Supplementary Fig. S2E). Further, seviteronel inhibited CD44^Hi^ tumorsphere formation across different TNBC cell lines in a dose-dependent manner, at doses below its IC50 (Supplementary Fig. S2F).

To test that the drug treatment effects observed were due to direct inhibition of AR, we transfected cells with a non-targeting control siRNA (siCT) or one of two independent siRNAs targeting AR (∼90% reduction in AR protein; Supplementary Fig. S2G). Again, we observed a significant reduction in the number and size of tumorspheres by siRNAs targeting exon1 or exon7 of AR (Fig. 2G-H). Importantly, this reduction was similar to that achieved by treatment with seviteronel, the most potent anti-androgen at inhibiting tumorsphere formation (Fig 2F).

To demonstrate that the anti-androgenic drugs were impacting an AR signalling network in basal CD44^Hi^ cells, we treated cells with vehicle, seviteronel, abiraterone or enzalutamide (10uM) for 48 hrs, and in parallel with siCT or siAR, followed by RNA-seq. Comparative analysis of resulting transcriptomes revealed gene regulatory networks altered by the different treatment conditions. There was a marked overlap across distinct AR pathways from the Molecular Signatures Database (MSigDB) (Fig. 2I-J; Supplementary Fig. S2H).

Compared to the other anti-androgens, seviteronel showed the strongest overlap with siAR in regulation of AR-associated pathways (Fig 2K). Gene set enrichment analysis (GSEA) revealed the mutual regulation of extracellular matrix components, collagen remodelling and TGF-β signalling by siAR and seviteronel, whereas abiraterone and enzalutamide commonly enriched for pathways associated with cell cycle and translation regulation (Fig. 2K; Supplementary Fig. S2I).

Together with the demonstration that seviteronel is the most effective anti-androgenic drug to inhibit CD44^Hi^ cancer stem cell function (Fig. 2E-F), these transcriptional data support the use of seviteronel to inhibit AR-dependent gene regulatory networks in basal CD44^Hi^ breast cancer cells.

### Inhibiting AR signalling prevents chemotherapy-induced cancer cell plasticity

We have shown that basal CD44^Lo^ cancer cells spontaneously convert to a CD44^Hi^ cancer stem cell state *in vitro* and *in vivo*, in a ZEB1-dependent manner ^14^. Consistent with further studies demonstrating that chemotherapy drives phenotypic plasticity ^16, 36^, we show that all standard chemotherapies used to treat TNBC (Doxorubicin, Cisplatin and Docetaxel) drive CD44^Lo^ to CD44^Hi^ cell state transitions in a dose dependent manner (Fig. 3A; Supplementary Fig. S3A). Importantly, both AR and ZEB1 were upregulated in a correlative manner as cells transition into the CD44^Hi^ state in response to chemotherapy (Fig. 3B-C).

**Figure 3:**
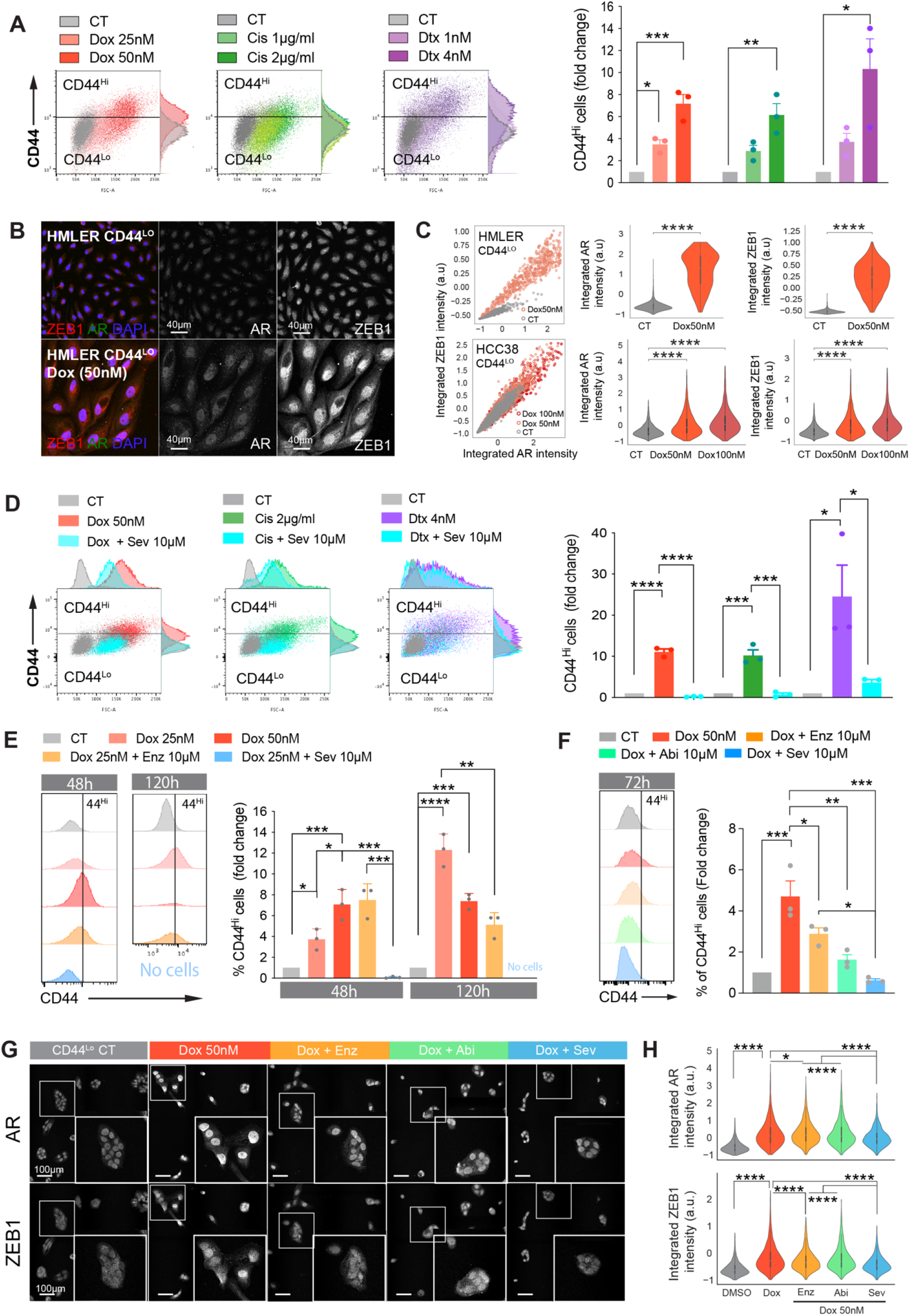
AR inhibition prevents chemotherapy-induced cancer cell plasticity. **A**, FACS analysis of dose-dependent induction of CD44^Lo^ to CD44^Hi^ cell state switching by doxorubicin (Dox), cisplatin (Cis) and docetaxel (Dtx) in HMLER cells. Quantification of newly-generated CD44^Hi^ cells presented in bargraph, right. **B**, Immunofluorescence for AR (green), ZEB1 (red) and DAPI (blue) in HMLER CD44^Lo^ cells treated with DMSO or Dox. **C**, Correlation and quantification of changes in AR and ZEB1 expression in HMLER (top) and HCC38 (bottom) CD44^Lo^ cells in response to Dox. **D**, FACS analysis of CD44^Lo^ to CD44^Hi^ cell state switching in HMLER CD44^Lo^ cells treated for 72h with DMSO (CT), Dox, Dox + Sev, Cis, Cis + Sev, Dtx, Dtx + Sev. Quantification of newly-generated CD44^Hi^ cells presented in bargraph, right. **E**, FACS analysis of HMLER CD44^Lo^ cells at 48h and 120h following treatment with DMSO, Dox or Dox + Sev showing switching to the CD44^Hi^ state (histograms, left) and quantification (bar plots, right). **F**. FACS analysis of HCC38 CD44^Lo^ cells at 72h following treatment with DMSO, Dox, or Dox plus Sev, Abi or Enz, showing switching to the CD44^Hi^ state (histograms, left) and quantification (bar plot, right). **G**, Immunofluorescence images of HCC38 CD44^Lo^ cells treated with DMSO, Dox, Dox + Enz, Dox + Abi or Dox + Sev showing AR and ZEB1. Boxes highlight magnified areas. **H**, Quantification of total ZEB1 and AR protein levels per cell in G.

To test the hypothesis that anti-androgen therapies prevents chemotherapy-induced cell state switching, we purified CD44^Lo^ cells and treated them with DMSO control (CT), chemotherapy alone, seviteronel alone, or the combination of seviteronel plus chemotherapy. Remarkably, the addition of seviteronel significantly inhibited chemotherapy-induced basal CD44^Lo^ cell state-switching (Fig. 3D; Supplementary Fig. S3B). Importantly, seviteronel was more effective than enzalutamide or abiraterone at preventing chemotherapy-induced cell state switching over multiple time points (48h, 72h, 120h and 144h) and various doses of different chemotherapies (Doxorubicin 25-50 nM, Cisplatin – 1-2 μg/ml, Docetaxel 1-4 nM) (Fig. 3E-F; Supplementary Fig. S3B,D). Of note, enzalutamide and seviteronel alone decreased CD44^Lo^ status compared to CD44^Lo^ cells treated with DMSO control, where seviteronel elicited a significantly larger decrease (Supplementary Figure S3C). In addition, seviteronel was also more effective than abiraterone or enzalutamide at preventing chemotherapy-induced upregulation of AR and ZEB1 (Fig. 3G-H; Supplementary Fig. 3H-I).

Together, these data suggest that combining seviteronel with chemotherapy could block *de novo* induction of chemotherapy-resistant CD44^Hi^ stem cells and may thereby provide a strategy to prevent the development of chemotherapy-resistant TNBC.

### Combining seviteronel with chemotherapy improves the effectiveness of chemotherapy in pre-clinical TNBC models

To determine the impact of anti-androgenic drugs alone or in combination with chemotherapy on primary and metastatic tumour growth, we implanted CD44^Hi^ MDA-MB-231-EGFP-Luc cells orthotopically into NOD/SCID mice. IHC analysis of primary tumours xenografts and matching metastases showed cytoplasmic AR enrichment in the cancer cells. (Fig. 4A). When primary tumours reached 100 mm^3^, animals were randomised into 8 different treatment groups: vehicle (Veh), an AR inhibitor alone (seviteronel; Sev, abiraterone; Abi, enzalutamide; Enz), chemotherapy alone (Docetaxel; Dtx), or chemotherapy plus an AR inhibitor (Dtx + Sev, Dtx + Abi, Dtx + Enz) and treated for two cycles (Fig. 4B). This experiment had two arms: Arm A where the primary tumour was present throughout the experiment and Arm B where the primary tumour was resected at the completion of Cycle 1.

**Figure 4:**
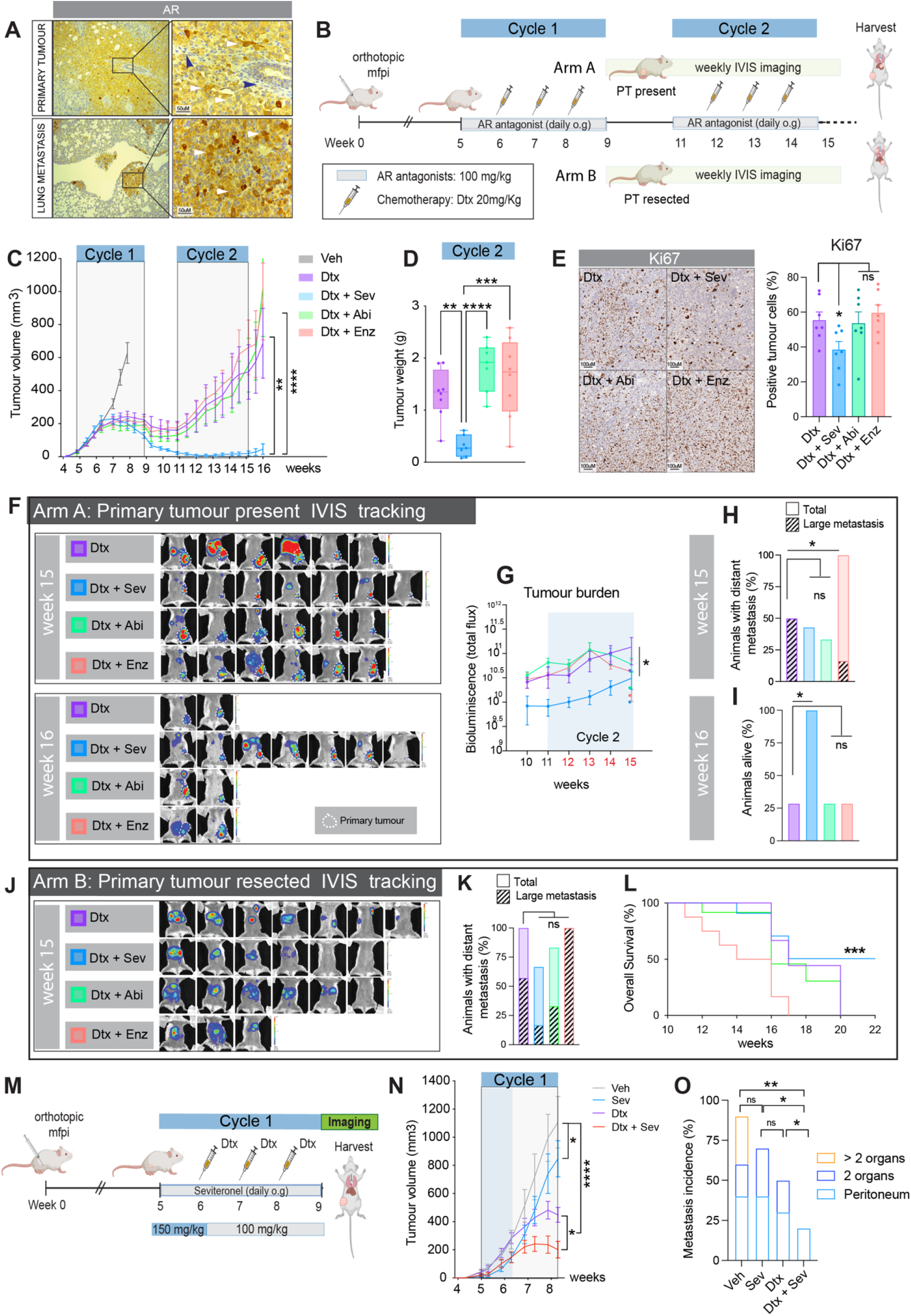
Combining seviteronel with chemotherapy reduces metastatic incidence and acquisition of chemotherapy-resistance in pre-clinical TNBC models. **A**, Immunohistochemistry (IHC) of AR staining in primary and lung metastasis from MDA-MB-231 xenograft model (left) and corresponding tissue magnification (right). Blue arrows: stroma cells. White arrows: cytoplasmic-AR+ cancer cells. **B**, Schematic of in vivo experimental design related to C-L. **C**, Growth-kinetics of MDA-MB-231 mice treated with Vehicle (Veh), 20 mg/kg Docetaxel (Dtx) or combination therapy (20 mg/kg Dtx + 100mg/kg/day Sev, Abi or Enz (n = 15 mice/group). **D**, Final tumour weights of animals harvested in Arm B at end of cycle 1. **E**, IHC staining and quantification for proliferation marker Ki67 in Dtx, Dtx + Sev, Dtx + Abi or Dtx + Enz in Arm B tumours harvested at end of cycle 1. **F**, IVIS bioluminescent images of luciferase in Arm A animals (primary tumour present) showing tumour burden at week 15 and 16. **G**, Quantification of bioluminescence (Total flux) from weeks 10-15 of animals in Arm A. **H**, Metastatic incidence (%) based on total flux in Arm A animals at week 15 (7 mice/group). Hashed portion of bar graph shows percentage of animals with high metastatic burden (measured as flux>10^10^). **I**, Incidence of Arm A animals alive at week 16. **J**, IVIS bioluminescent images of luciferase in Arm B animals (primary tumour resected) showing tumour burden at week 15 (7 mice/group). **K**, Metastatic incidence (%) based on total flux in Arm B animals at week 15. Hashed portion of bar graph shows percentage of animals with high metastatic burden (measured as flux>10^10^). **L**, Kaplan-Meier survival curve of Arm B animals. **M**, Schematic of in vivo experimental design related to N-O. **N**, Growth kinetics from animals treated with Veh, Sev, Dtx or Dtx + Sev (n = 12 mice/group). **O**, IVIS quantification of bioluminescence representing metastatic burden in animals from N.

We observed no significant inhibition of primary tumour growth in animals treated with an AR inhibitor alone compared to vehicle. Those 4 treatment groups reached ethical end-point at the end of Cycle 1 and were thus euthanised and tumours harvested (Supplementary Fig. S4A-S4C). As expected, chemotherapy alone significantly reduced primary tumour volume at the completion of Cycle 1 (Supplementary Fig. S4B-S4C), however, tumours became chemotherapy-resistant and grew exponentially in Cycle 2 (Fig. 4C). The addition of abiraterone or enzalutamide to docetaxel treatment did not alter primary tumour growth compared to chemotherapy alone in any cycle (Fig. 4C). In contrast, the combination of seviteronel and docetaxel markedly reduced primary tumour growth compared to docetaxel alone in Cycle 2 (Fig. 4C-D).

Markers of proliferation (Ki67) and apoptosis (CASP3) were assessed in the primary tumours resected at the end of cycle 1 (Arm B). Ki67 staining was significantly decreased in the Dtx + Sev group compared to Dtx alone (Fig. 4E). While no significant differences were observed in CASP3 staining across the tumour groups (Supplementary Fig. S4E). These data indicate that the anti-androgen seviteronel prevented the emergence of chemotherapy-resistant disease in a manner that the other anti-androgenic drugs did not (Fig. 4C-D; Supplementary Fig. S4D-S4E).

Next, we determined the impact of combination anti-androgen and chemotherapy treatments on metastatic development (via IVIS tracking) on mice bearing primary tumours throughout the experiment (Arm A, Fig. 4F). Total tumour burden (primary tumour plus metastases) was significantly lower only in the seviteronel plus docetaxel combination treatment group compared to the chemotherapy alone (Fig. 4G). Indeed, at 16 weeks, 100% of animals treated with seviteronel plus docetaxel were alive after completing Cycle 2 compared to only 25% of animals for all other groups (Fig. 4I). While we did observe a reduction in the number of large metastases (>10^10^ total flux) in the abiraterone plus Docetaxel combination treatment group compared to the chemotherapy alone (Fig. 4H), this did not confer greater survival (Fig. 4I). Surprisingly, when enzalutamide was combined with chemotherapy, there was a significant increase in metastases throughout the body (Fig. 4H). This unexpected result raises concern about use of this anti-androgenic drug to treat women with TNBC.

We next analysed metastatic development in animals following primary tumour resection at the end of cycle 1 (Fig. 4J), an experimental design that mimics removal of primary tumours shortly after neo-adjuvant treatment in the clinical setting (Arm B, Fig. 4B). IVIS tracking was performed weekly (Supplementary Fig. S4F). Again, we observed that seviteronel plus docetaxel reduced metastatic tumour burden compared to the chemotherapy alone, with 28.5% of animals remaining metastasis-free at the conclusion of the experiment (Fig. 4K; Supplementary Fig. S4F). This corresponded to a significant improvement in overall survival compared to chemotherapy alone (Fig. 4L). No significant benefit was observed for the combination of abiraterone and chemotherapy (Fig. 4J-L). Consistent with the increase in tumour burden observed when enzalutamide was combined with chemotherapy in Arm A (Fig. 4H), in Arm B this combination treatment also had a significant increase in metastatic burden compared to the chemotherapy alone (determined by >10^10^ total flux, Fig. 4K) and led to a decrease in overall survival (Fig. 4L).

Given that Seviteronel outperformed Abiraterone and Enzalutamide when used in combination with Docetaxel *in vivo*, we increased the dose of Seviteronel from 100 mg/kg/day to 150mg/kg/day to test if the reduction in metastasis and increase in survival could be further enhanced (Fig. 4M). However, the higher dose was not well tolerated and after the first week of treatment we reduced it back to 100 mg/kg/day (as per early treatment regimen, Fig. 4B) for the remainder of the experiment. Interestingly, short exposure to the higher Seviteronel dose showed a small but significant reduction in tumour growth with the anti-androgen as a single-agent, while the combination of this drug with chemotherapy significantly reduced primary tumour volume compared to chemotherapy alone (Fig. 4N). Moreover, we observed a decrease in metastatic incidence and in the number of organs seeded for chemotherapy alone and Seviteronel alone, but even more striking was again the absence of multi-organ metastatic lesions for the Seviteronel plus chemotherapy combination (Fig. 4O).

### Seviteronel pairs with distinct chemotherapies to improve targeting of advanced TNBC patient-derived xenograft models

To assess the effectiveness of combining seviteronel and chemotherapy in clinically relevant models of advanced TNBC, we first selected a TNBC patient-derived xenograft (PDX), Gar12-58, derived from a liver metastasis biopsy. This PDX model harboured a mix of CD44^Lo^ and CD44^Hi^ cell populations (Fig. 5A). Corresponding with this profile, low and high cytoplasmic AR levels with scattered positive nuclei were detected, as well as scattered cells showing nuclear ZEB1 positivity (Fig. 5A). Using this model, we conducted experiments with 1 treatment cycle, where following an initial period of tumour engraftment, animals were randomised to treatment with seviteronel alone or in combination with Docetaxel, Cisplatin or pegylated-doxorubicin (Fig. 5C-E, Supplementary Fig. S5D). Echoing earlier results with the cell line xenografts, seviteronel as a single agent (100 mg/kg/day) did not reduce primary PDX tumour volume compared to vehicle control (Fig. 5C, and Supplementary Fig. S5A-D), but the combination of seviteronel and docetaxel significantly reduced tumour growth after completion of treatment (Supplementary Fig. S5C) and delayed regrowth of tumours post-chemotherapy (Fig. 5C). Accordingly, the combination treatment significantly improved overall survival compared to chemotherapy alone (Fig. 5D).

**Figure 5:**
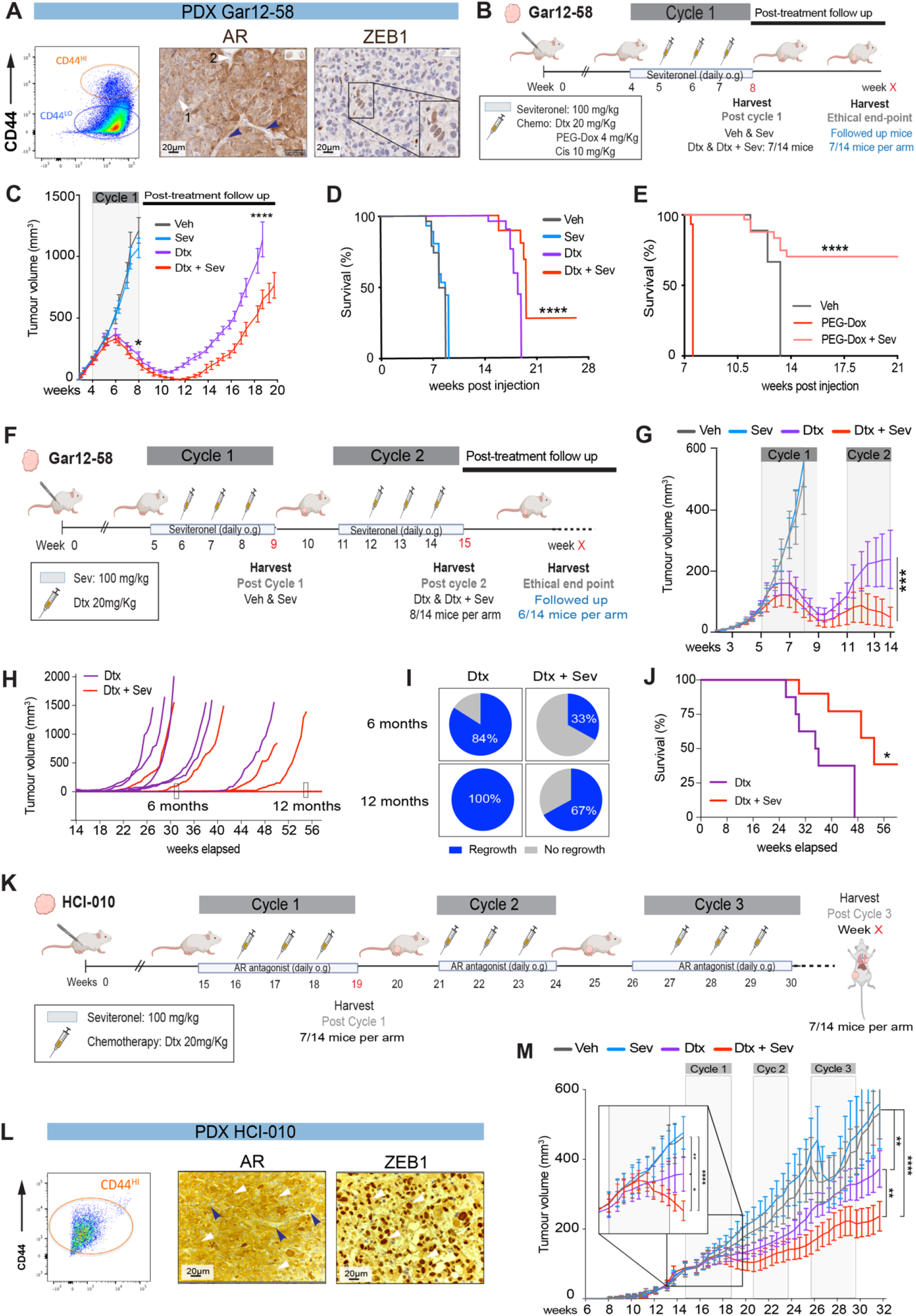
Seviteronel pairs with distinct chemotherapies to improve targeting of advanced TNBC patient-derived xenograft models. **A**, FACS profile showing CD44 status and immunohistochemistry (IHC) of AR or ZEB1 staining in the Gar12-58 triple-negative breast cancer patient-derived xenograft (PDX) model. White arrows indicate cancer cell with low (1) and high (2) cytoplasmic AR levels. Blue arrows highlight stroma. **B**, Schematic of in vivo experimental design related to C-E. **C**, Growth-kinetics of Gar12-58 PDX treated with vehicle (Veh), 20 mg/kg docetaxel (Dtx), 100 mg/kg/day seviteronel (Sev) or 20 mg/kg Dtx + 100 mg/kg/day Sev (n = 14 mice/group). **D**, Kaplan-Meier survival analysis of animals in C. **E**, Kaplan-Meier analysis of Gar12-58 PDX treated with Veh, 4 mg/kg Pegylated-Doxorubicin (PEG-Dox) or 4 mg/kg PEG-Dox + 100 mg/kg/day Sev. **F**, Schematic of in vivo experimental design related to G-I. **G**, Growth kinetics of Gar12-58 PDX treated with Vehicle (Veh), 100 mg/kg/day Sev, 20 mg/kg Dtx or 20 mg/kg Dtx + 100 mg/kg/day Sev (n = 15 mice/group). **H**, Post-cycle 2 (15w onwards) growth kinetics of animals previously treated with Dtx or Dtx + Sev (in G). **I**, Summary of primary tumour regrowth incidence in animals from H. **J**, Kaplan-Meier survival analysis of animals from H. **K**, Schematic of in vivo experimental design related to L-M. **L**, FACS profile showing CD44 status and immunohistochemistry (IHC) of AR (white arrows indicate cytoplasmic and nuclear positive cancer cells. Blue arrows highlight stroma) and ZEB1 staining in the HCI-010 triple-negative PDX model (white arrows indicate examples of strong nuclear ZEB1 positive cancer cells). **M**, Growth kinetics of animals from K treated with Veh, Sev, Dtx or Dtx + Sev along 3 treatment cycles (n = 14 mice/group).

At the dose used, Cisplatin (10 mg/kg) alone or in combination with seviteronel did not reduce the growth of Gar12-58 PDX tumours (Supplementary Fig. S5A). However, we observed striking effects on tumour growth and survival by PEG-Dox (4mg/kg) alone and in combination with seviteronel. While Peg-Dox alone significantly reduced tumour growth compared to vehicle (Supplementary Fig. S5B), animals died due to toxicity much earlier than vehicle controls (Fig. 5E). However, the addition of seviteronel rescued animals treated with PEG-Dox, rendering this form of chemotherapy tolerable for the animals (Fig. 5E). Moreover, the combination produced a robust and sustained inhibition of primary tumour growth, with a concomitant increase in body weight that led to a remarkable overall survival benefit (Fig. 5E; Supplementary Fig. S5C).

Collectively, these data demonstrate that the cytotoxic effects of chemotherapies that inhibit tumour growth alone, and thus, overall survival, can be significantly improved by the addition of seviteronel.

In a subsequent experiment, we aimed to determine if survival of animals bearing Gar12-58 PDX tumours could be further enhanced by additional treatment cycles of combination seviteronel plus docetaxel (Fig. 5F). For this experiment, half of the animals treated with Dtx and Dtx + Sev were harvested at the completion of Cycle 2 for analysis, and, the remaining animals continued to be monitored for primary tumour growth post-cycle 2 treatment. In the cohort harvested at completion of the two cycles, primary tumours were significantly smaller in the Seviteronel plus Docetaxel group compared to the chemotherapy alone (Fig. 5G and Supplementary Fig. S5E). In the second cohort of animals monitored for post-treatment survival, tumours regrew in 100% of animals in the Docetaxel arm 12 months post-commencing treatment, while tumours remained undetected in 33% of animals in the combination treatment arm (Fig. 5H–I). Thus, seviteronel plus Docetaxel significantly increased overall survival compared to Docetaxel treatment alone (Fig. 5J).

Next we used a TNBC PDX model (HCI-010) derived from a lung metastasis of a patient heavily treated with chemotherapy. Consistent with chemotherapy-mediated selection or induction of cancer stem cells, this PDX model had a CD44^Hi^ enriched cancer cell profile (Fig. 5L). Cancer cells in this model expressed AR in the cytoplasm and nucleus, while ZEB1 exhibited strong nuclear expression (Fig 5L). Following a 3-cycle treatment regimen (Fig. 5K), seviteronel treatment alone (100 mg/kg/day) did not significantly reduce primary HCI-010 tumour growth compared to vehicle (Fig. 5L). However, seviteronel combined with Docetaxel significantly reduced primary tumour growth compared to the chemotherapy alone following the first cycle of treatment (Fig. 5L). This effect was further enhanced at the conclusion of the treatment cycles (Fig. 5L; Supplementary Fig. S5F).

Together, these *in vivo* experiments demonstrate that the addition of seviteronel increased the therapeutic efficacy of chemotherapy alone and prevents the emergence of chemotherapy-resistant disease, thereby improving overall survival in cell line xenograft and PDX models of TNBC.

### Cytoplasmic AR expression is prognostic and predicts poor response to chemotherapy in TNBC patient cohorts

High cytoplasmic expression of AR was notable in all *in vivo* TNBC models used in this study that contain CD44^Hi^ cells (Fig. 4A, Fig. 5A, K). Cytoplasmic AR however, is not scored in the clinical setting. Clinical identification of AR positive tumours is traditionally determined by calculating the H-score for nuclear expression only, where H-score is determined by the sum of [1 × (% cells 1+) + 2 × (% cells 2+) + 3 × (% cells 3+)] ^37^. To investigate the utility of cytoplasmic AR (Cyt-AR) as a prognostic marker for disease outcome in TNBC, we defined the H-score for Cyt-AR and nuclear AR (Nuc-AR) in 308 patient samples from two independent, treatment-naive TNBC cohorts and determined their relationship to survival outcome ^38, 39^.

AR staining was present in the nucleus or cytoplasm (H-score>0) in 25% of treatment-naive TNBC tumours (Fig. 6A–6B, Supplementary Fig. S6A). A Cox Proportional Hazard model demonstrated that Nuc-AR is not significantly associated with good or bad prognosis in treatment-naive TNBC patients (HR=0.72, p=0.228) (Fig. 6C-D). However, Cyt-AR is prognostic for poor overall survival in treatment-naive TNBC patients (HR=1.94; p=0.029; Fig. 6C). Accordingly, tumours with high Cyt-AR were associated with a 19-month reduction in overall patient survival benefit compared to patients with low Cyt-AR tumours (36 months and 55 months, respectively; p=0.014; Fig. 6D).

**Fig 6.**
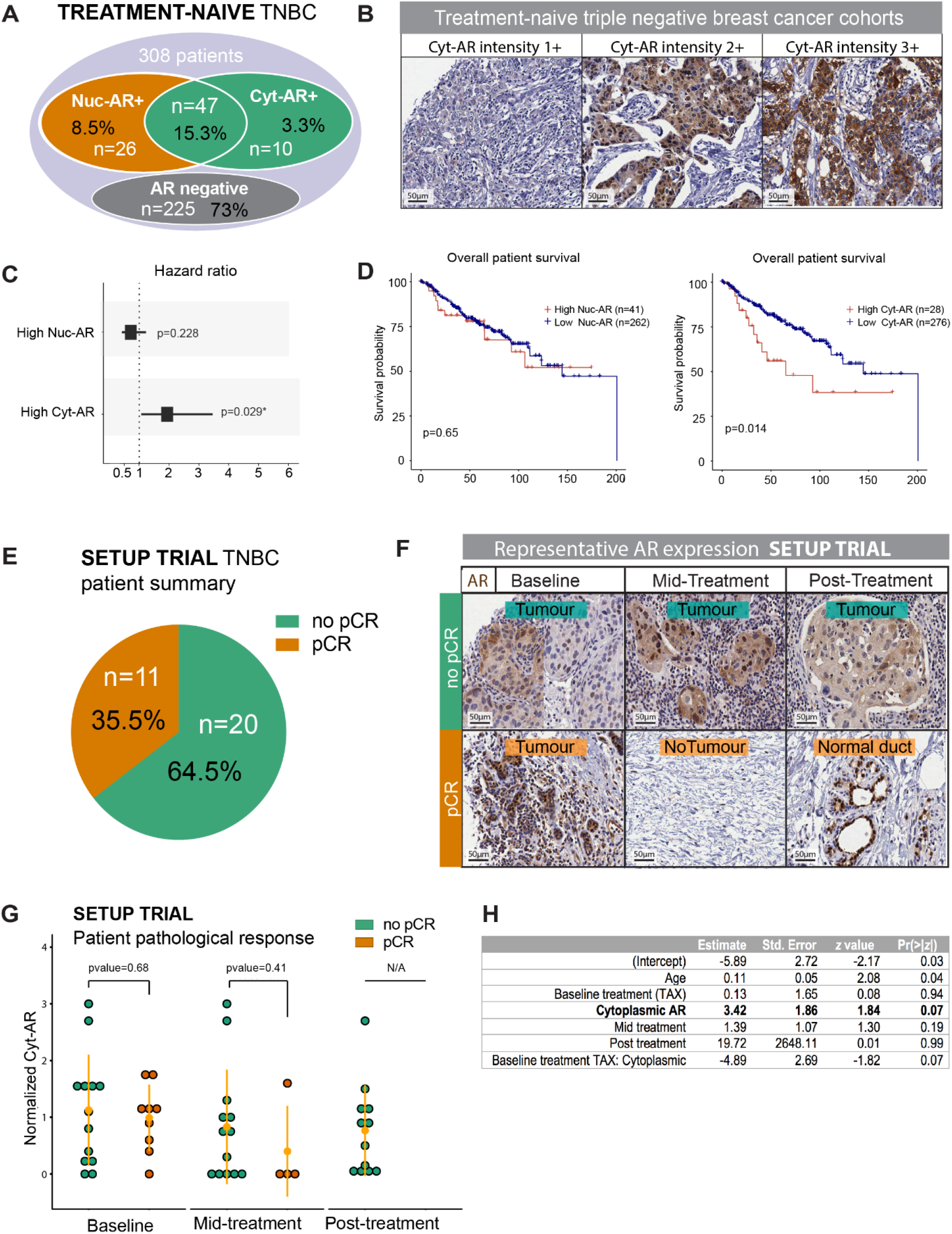
Cytoplasmic AR expression is prognostic and predicts poor response to chemotherapy in clinical TNBC cohorts. **A**, Venn diagram displaying representation of AR staining in two independent treatment-naïve TNBC patient cohorts. **B**, Representative IHC images of AR staining from A. **C**, Hazard ratios from Cox Proportional Hazard models on prognostic power of nuclear (Nuc) and cytoplasmic (Cyt) AR staining from B. **D**, Kaplan-Meier plots assessing survival based on nuclear or cytoplasmic expression in patients from B. **E**, Summary of patient data from SETUP trial reflecting pathological complete response (pCR) outcome. **F**, Venn diagram displaying representation of AR staining in SETUP trial patients from E. **G**, Representative IHC images of AR staining from E. **H**, Analysis of cytoplasmic AR (Cyt-AR) in SETUP trial patients and relationship to pathological complete response. **I**, Logistic regression model assessing ability of Cyt-AR to predict poor response to chemotherapy in SETUP trial data.

Around 1/3 of TNBC patients do not obtain a pathological complete response (pCR) to their primary chemotherapy, which indicates that disease is likely to recur ^20^. To date, there is no way to identify which TNBC patients at diagnosis will reach a pCR following primary chemotherapy. Having demonstrated that Cyt-AR is prognostic for poor outcome in treatment-naive TNBC (Fig. 6C-D), we wanted to determine if Cyt-AR predicts poor response to chemotherapy. To do this, we analysed Cyt-AR and Nuc-AR expression in a cohort of 29 locally advanced TNBC patients with FFPE samples collected pre-treatment (Baseline), mid-treatment (Mid) and post-chemotherapy treatment (Post) ^40.^ In that study, 31 patients were randomised to receive four cycles of anthracycline-based chemotherapy (FEC), followed by four cycles of Docetaxel therapy (Arm A), or the same regimen in reverse order (Arm B) ^40^. Over a third of patients (11/ 31; 35%) obtained a pCR, while the remainder (20/31; 65%) did not (Fig. 6E, Supplementary Fig. S6A). We stained samples for AR and determined the H-score for Cyt-AR and Nuc-AR (Fig. 6F-6G; Supplementary Fig. S6B) and discovered that Cyt-AR was present in 20% of this patient cohort at baseline. Using a logistic regression model, we determined that baseline Cyt-AR was predictive of no pCR to chemotherapy (p=0.0663). Given the small sample size, we considered a 10% Type 1 error (p<0.1) significant for this dataset (Fig. 6H).

Together, these data demonstrate that baseline Cyt-AR expression, and not Nuc-AR expression, identifies poor prognosis TNBC patients and is predictive of patients that do not obtain a pCR to standard-of-care chemotherapy.

### Seviteronel treatment followed by chemotherapy is associated with improved overall survival in cytoplasmic AR-positive TNBC patients

Seviteronel was tested as a monotherapy in the Clarity-01 Phase II Clinical trial (NCT02580448) for safety and efficacy in patients with advanced-stage breast cancer ^41^. On average, patients were treated with Seviteronel for a median of 56 days and the treatment was proven safe. Based on our observations that Cyt-AR is associated with poor prognosis in TNBC (Fig. 6), we performed a post-hoc biomarker analysis to evaluate whether Cyt-AR H-scores were associated with improved overall survival in TNBC patients treated with seviteronel in the Clarity-01 trial.

We evaluated Cyt-AR expression in tumours from 89 TNBC patients treated with seviteronel and observed a trend for improved overall survival in Cyt-AR high patients (H-score >/=100) compared to Cyt-AR low patients (H-score <100) (HR=0.71; p=0.24) with an 8-week improvement in median overall survival from 49 to 57 weeks (Fig. 7A, and Supplementary Fig. S7A). This contrasted with our observation that high Cyt-AR is associated with poor prognosis in treatment-naïve TNBC (Fig. 6).

**Figure 7:**
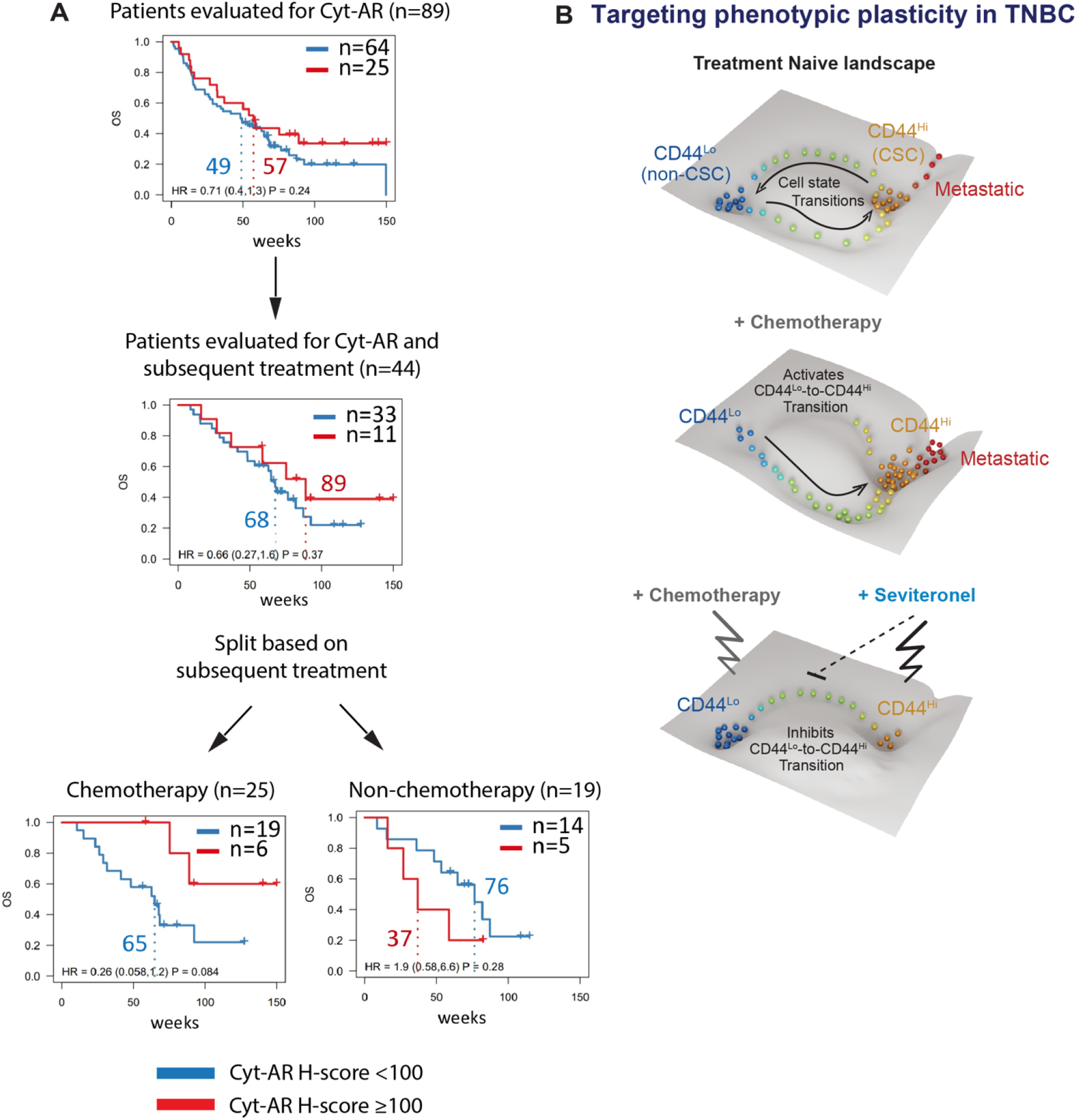
Seviteronel treatment followed by chemotherapy is associated with improved outcome in cytoplasmic AR+ TNBC. **A**, Overall survival analysis of the Phase II Clarity-01 clinical trial. Kaplan-Meier plots for overall Survival (OS) in patients treated with seviteronel monotherapy split into low and high cytoplasmic AR (Cyt-AR) expression based on H-score < 100 or ≥ 100. HR, hazard ratio with 95% confidence interval in brackets. Dotted lines indicate median survival times with 95% confidence interval in brackets. **B**, Targeting phenotypic plasticity as a new therapeutic strategy for cancer. Cancer cells reside in distinct phenotypic states upon a cell state landscape. Treatment with chemotherapy forces cells to move across the landscape and switch from a CD44^Lo^ chemotherapy-sensitive cell state into an aggressive CD44^Hi^ chemotherapy-resistant cell state. Strategies to prevent cell state switching in response to chemotherapy and/or to push aggressive cancer cells into a chemo-sensitive state will translate into effective therapies in the clinic. We show that inhibiting AR signalling with Seviteronel has a dual function: 1) it blocks the aggressiveness of the CD44^Hi^ cell state, and 2) it prevents CD44^Lo^ to CD44^Hi^ cell state switches to maintain cells in the CD44^Lo^ chemotherapy-sensitive cell state. Hence, the combination of seviteronel plus chemotherapy eradicates cancer cells better than chemotherapy alone.

Follow-up data detailing subsequent treatments after seviteronel cessation was available for 44 out of the 89 patients (Supplementary Fig. S7B). In this patient subset, tumours with high Cyt-AR high were associated with a similar trend for improved overall patient survival compared to tumours with low Cyt-AR (HR=0.66; p=0.37), with a 21-week improvement in median overall patient survival from 68 to 89 weeks (Fig. 7A; Supplementary Fig. 7A-B).

Given that our pre-clinical work demonstrated that seviteronel sensitises tumours to chemotherapy, we further sub-grouped the 44 patients according to the type of therapy received following seviteronel: those that received chemotherapy (n=25), versus those that received a non-chemotherapeutic treatment (n=19) (Supplementary Fig. S7B). Marked differences in these patient populations were observed. In the non-chemotherapy subgroup, patients with high Cyt-AR tumours fared worse than patients with low Cyt-AR tumours (HR=1.9; p=0.28) with a median overall survival of 37 and 76 weeks, respectively. These findings are in line with our earlier data demonstrating that tumours with high Cyt-AR are associated with poorer patient prognosis compared to tumours with low Cyt-AR (Fig. 7A-7D) and suggest that seviteronel did not improve survival outcomes in that patient population.

In the chemotherapy group, tumours with low Cyt-AR were associated with a median overall patient survival of 65 weeks. Remarkably, however, median overall survival was not reached in patients with tumours that had high Cyt-AR expression, as 2/3rds of patients were still alive at the study endpoint (HR=0.26; p=0.084; Fig. 7A, Supp Fig. 7A-B). In line with our pre-clinical data, these results suggest that patients received a meaningful clinical benefit from pairing seviteronel with chemotherapy.

Comparing tumours with different TNBC molecular subtypes on this trial (defined using Nanostring data), we observed that median Cyt-AR expression was higher in LAR compared to other subtypes, however high Cyt-AR (H ≥ 100) was evident across all subtypes (Supplementary Fig. S7C). Thus, Cyt-AR was not identifying response in patients of a particular TNBC subtype (Supplementary Fig. S7D). These data suggest high Cyt-AR is an independent indicator of patient response to this treatment combination

Similar to the treatment-naïve TNBC cohort (Fig. 7D), Nuc-AR expression in the primary tumour did not associate with an overall survival benefit in the Seviteronel followed by chemotherapy or non-chemotherapy treated subgroups (Supplementary Fig. S7E-F).

Collectively, these clinical data suggest Cyt-AR expression identifies a group of TNBC patients that received an overall survival benefit when treated sequentially with seviteronel followed by chemotherapy.

## DISCUSSION

There is an unmet need to improve survival outcomes for patients for which there are currently no effective therapies ^12^. Targeting phenotypic plasticity holds much promise as a novel therapeutic strategy to treat those patients. Towards that goal, we now demonstrate that targeted inhibition of phenotypic plasticity in combination with standard-of-care chemotherapies, prevents the emergence of chemotherapy-resistant disease.

In the current study, we identify a unique AR signalling network that plays a dual role in TNBC: it is critical for maintaining the function of aggressive CD44^Hi^ cancer stem cells, and it is a critical mediator of CD44^Lo^ to CD44^Hi^ phenotypic plasticity. To find a means of inhibiting AR signalling in this setting, we tested three clinically available anti-androgenic drugs (enzalutamide, abiraterone, seviteronel) and showed that seviteronel was the most efficacious at inhibiting the function of aggressive CD44^Hi^ cancer stem cells and preventing spontaneous and chemotherapy-induced *de novo* generation of CD44^Hi^ cancer stem cells *in vitro*. Of note, we show that AR mediates these effects in part by modulating ZEB1 expression. Moreover, we demonstrate in pre-clinical cell line and patient-derived xenograft models that seviteronel in combination with chemotherapy inhibits primary and metastatic tumour growth significantly better than chemotherapy alone, which translates into increased overall survival. These data demonstrate that targeting existing cancer stem cell populations and simultaneously preventing the emergence of new cancer stem cells may be a transformative therapy for cancer patients, especially those that are at risk of poor response to primary chemotherapies. The current landscape of targeted therapies for TNBC patients offers little hope.

Therapeutic targets are available for some patients with metastatic TNBC, including PARP inhibitors for patients with germline BRCA1 and BRCA2 mutations that may convey sensitivity to this therapeutic strategy ^42^. In clinical studies for patients receiving PARP inhibitors, a gain in progression-free survival (PFS) of 7 months compared to 4 months for standard-of-care chemotherapy was obtained, with no change in overall survival ^42^. In other clinical studies, expression of human trophoblast cell-surface antigen 2 (Trop-2) receptor could enable sensitivity to the antibody-drug conjugate sacituzumab-govitecan ^43^, offering patients an overall survival benefit of 12.1 months compared to 6.7 months with standard chemotherapy. Although these options are promising, there remains an urgent need for development of more effective targeted therapies that improve overall survival for treatment-refractory TNBC. In the past decade, there has been great interest in targeting the AR for this disease sub-type ^30, 31, 34, 44^, but no compelling rationale for what strategy is best and no development of biomarkers to predict treatment response.

The lack of biomarkers to distinguish patients that will respond well to primary chemotherapy treatments from those that will develop chemotherapy-resistant disease is a significant issue for oncologists and TNBC patients. The ability to do so would serve two purposes: 1) it would enable patients undergoing primary treatment to know that their cancer is unlikely to recur, and 2) it would identify patients at diagnosis that are highly likely to need immediate aggressive and ideally, complementary targeted therapies to curb their disease. To address that knowledge-gap, we show that cytoplasmic, not nuclear, AR expression identified treatment-naive patients with poor overall survival. Furthermore, in a TNBC patient cohort that provided pre-, mid- and post-chemotherapy biopsies, we show that cytoplasmic AR expression was associated with lack of pCR to standard-of-care chemotherapy. Together, our data demonstrate that standard IHC analysis of cytoplasmic AR expression is a potential biomarker that has the power to determine at diagnosis, patients at high risk of developing chemotherapy-resistant disease. Based on these findings, and given that AR expression has traditionally been clinically scored only for nuclear content, we propose that incorporating pathological scoring of nuclear and cytoplasmic AR expression in all breast cancer biopsies should become routine procedures alongside ER, PR and HER2. Moreover, TNBC patients whose tumours express cytoplasmic AR would be candidates for cancer cell plasticity therapies, such as our newly-defined combination treatment with seviteronel plus chemotherapy. Whether this may be efficacious in other forms of disease, such as tamoxifen resistant ER+ breast cancer where cytoplasmic AR may promote growth ^45^, remains to be determined.

In addition to their CYP17-lyase inhibitor function to reduce androgen levels, seviteronel and abiraterone also act as direct AR antagonists that displace AR ligands from the AR ligand binding domain ^46.^ Importantly, seviteronel is as effective as enzalutamide at inhibiting induction of AR target genes and recruitment of AR to target promoters ^46, 47^. Together, its molecular properties may account for the greater concordance in regulation of AR target genes observed between siAR and seviteronel treatment compared to either abiraterone or enzalutamide, and may in part account for the increased efficacy of seviteronel over those inhibitors when used in combination with chemotherapy *in vivo*.

Anti-androgenic drugs (abiraterone acetate, bicalutamide, enzalutamide) have been assessed as single agents in nuclear AR+ TNBCs with minimal success ^30, 31, 34, 44^. In single-arm Phase II clinical trials, the median PFS for abiraterone acetate was 11 weeks, PFS for bicalutamide was 12 weeks, and PFS for enzalutamide was 12.6 weeks. No benefits in overall survival (OS) were observed for those agents. While no PFS or OS benefit was observed for single agent seviteronel in a similar Phase II study design ^41,^ our post-hoc biomarker analysis demonstrated that patients with high Cyt-AR positive tumours, corresponding to highly plastic/CSC-enriched tumours, received a remarkable overall survival benefit from treatment with seviteronel followed by subsequent chemotherapy treatment.

Together, our data demonstrate that seviteronel combined with chemotherapy could be an effective therapeutic strategy for TNBC, acting in a manner similar to combining trastuzumab (Herceptin) with chemotherapy for HER2 positive breast cancers ^48^. Furthermore, we have identified that cytoplasmic AR expression is a potential biomarker to predict patients most likely to respond to this novel therapeutic strategy. Our data suggest that cytoplasmic AR expression in primary tumours may mark plastic CD44^Lo^ cancer cells with the ability to transition to a more aggressive (high Cyt-AR), chemotherapy-resistant state (Fig. 7B). Moreover, we have determined that seviteronel is the most efficacious anti-androgen to combine with chemotherapy to achieve improved inhibition of primary and metastatic tumour growth. Accordingly, we have initiated a Phase I clinical trial to test the safety and efficacy of combining seviteronel with docetaxel chemotherapy for metastatic TNBC patients (NCT04947189). This clinical trial will determine the ability to target cancer stem cells, and prevent new ones from emerging, as a promising strategy for TNBC, and likely, other cancer types.

## Data Availability

All data produced in the present work are contained in the manuscript or will be made available upon reasonable request to the authors.

## ACKOWLEDGEMENTS

We are grateful to the core histology and flow cytometry facilities at the Garvan Institute of Medical Research. We thank all members of the Cancer Cell Plasticity lab for the constant discussion and development of this project. This work has been improved through the helpful discussion and feedback from colleagues including Elizabeth Williams, Rik Thompson, Jeff Holst, Tri Phan and Priyamvada Rai. We extend a huge thank you to our families for supporting us on this scientific journey: Paul, Gabrielle, Alice and Jonathan Bernath, and Gonzalo Pérez Siles.

## FUNDING

This research was supported by: St. Vincent’s Hospital Research Grant, NHMRC APP1088122 and APP1181230, National Breast Cancer Foundation research grant IIRS-19-092. L.R. is supported by a Ramon-Areces Fellowship. C.L.C and B.P.S.J were supported by the Nelune Foundation Rebecca Wilson Fellowship, C.L.C was supported by a CINSW Fellowship CDF181243. L.D.G. was supported by the Kinghorn Foundation. TEH was supported by a National Breast Cancer Foundation fellowship (IIRS-19-009). FVK and CAP are supported by an Australian Government Research Training Program (RTP) Scholarship.

## AUTHOR CONTRIBUTIONS

B.P.S.J and C.L.C conceived the project, B.P.S.J was involved in all experimental research done in this project, manuscript writing and review. S.H-Z performed computational analyses on RNA-seq and histological data, L.R, L.C, A.B, A.K conducted in vitro experiments, V.R performed in vitro and all animal experiments, H.H.M performed RNA-seq and computational analyses, C.A.P performed IHC quantitative analysis F.K analysed immunofluorescence experiments, D.D.B performed protein network analyses, E.L provided access to PDX models, V.G provided access to clinical samples, R.D provided clinical expertise on experimental design, access to clinical samples, and clinical data interpretation, C.V performed pathological scoring of AR on all clinical samples, S.O.T provided access to clinical samples, T.E.H provided expertise on AR and critical review of the manuscript, J.G.L performed IF analyses, contributed to manuscript writing and review, L.D.G analysed clinical data, contributed to manuscript writing and review, M.J.D oversaw computational analyses performed by S.H.Z and D.D.B and contributed to manuscript writing and review. C.L.C supervised project, provided funding, wrote manuscript.

## COMPETING INTERESTS

C.L.C is the founder and Managing Director of Kembi Therapeutics Pty Ltd that holds US patents for seviteronel. C.L.C and B.P.S.J are co-inventors on a patent for the method of use of seviteronel in combination with chemotherapy to treat cancer.

## DATA AVAILABILITY STATEMENT

The datasets generated during and/or analysed during the current study are available from the corresponding author on reasonable request, and/or have been uploaded to GEO omnibus, and/or will be made publicly available upon publication of the manuscript.

**NB48 onwards appear in methods and supplementary information only**

## METHODS

### Statistical analysis

#### In vitro studies

Due to non-normality of the single-cell quantitative immunofluorescence data, Wilcoxon Rank Sum Tests were performed to compare AR and ZEB1 single cell immunofluorescence distributions. Pearson’s correlation coefficient was used to measure correlation between ZEB1 and AR expression.

IC50s were determined using a Nonlin fit with a [Inhibitor] vs. normalized response – Variable slope model for drug response studies.

Ordinary one-way ANOVA applying the Tukey’s multiple comparison test was used to define statistical significance in tumorsphere assays, western blots and FACS analysis of treatment-induced plasticity.

#### In vivo studies

Two-way ANOVA multiple comparison test was applied when comparing growth kinetics for 4 different treatment arms. When comparing 2 treatment arms a paired t test was used instead.

One-way ANOVA Tukey’s multiple comparison test was applied to determine differences among tumour weights, tumour volume evolution in response to treatment and IHC Ki67/CASP3 staining scores.

Log-rank (Mantel-Cox) test was used to determine differences in survival between treatment arms.

One-way ANOVA Krustal-Wallis test was applied for survival and metastatic incidence analyses due to the small sample size analysed. Paired t test was used to compared differences in flux intensity between Dtx and Dtx + Sev treatments at the end of each treatment cycle.

The following statistical thresholds have been applied through the study: *p<0.05, **p<0.01, ***p<0.001 and **** p<0.0001.

Data is represented as mean +/-standard error (SE) for all analyses unless otherwise stated.

### Cell Culture

MCF7 (Michigan Cancer Foundation), ZR75-1, MDA-MB-231 (American Type Culture Collection) and the SUM159PT (Cellosaurus) cell lines were verified through short tandem repeat profiling and tested negative for mycoplasma contamination. Parental cell lines were cultured in DMEM, RPMI 1640 media, DMEM-F12 and Ham’s F-12 + 1% (v/v) hydrocortisone respectively (ThermoFisher). Media were supplemented with 10% (v/v) fetal bovine serum (GE Healthcare), 20 mM HEPES (ThermoFisher).

HMEC cells were purchased from ATCC and transformed into HMLER cells by the sequential addition of hTERT, SV40-ER and RAS as previously described^21^. Cells were cultured in serum-free mammary epithelial growth medium (MEGM) and MEGM Single Quots (Lonza, cat no. CC-4136: 1 ml bovine pituitary extract (BPE), 0.5 ml GA-1000 (30 mg/ml Gentamicin and 15 μg/ml Amphotericin), 0.5 ml insulin, 0.5 ml hydrocortisone and 0.5 ml hEGF). Cells were maintained at 37°C with 5% CO_2_. HCC38 cells were purchased from ATCC and cultured in RPMI containing 10% (v/v) FBS (PS; 5.000 units penicillin and 5 mg streptomycin/ml in H_2_O, Sigma Aldrich, cat no. P4333). All cell lines were routinely tested to confirm the absence of mycoplasma contamination. All cell line-specific media were supplemented with 1% (v/v) penicillin-streptomycin.

All in vitro and in vivo experiments were graphed and analysed using Prism9 software, unless otherwise specified.

### Drugs

Drugs were obtained from the following sources: Seviteronel (Innocrin Pharmaceuticals, North Carolina, USA), Enzalutamide (Selleck Chemicals, Houston, USA), Doxorubicin (Sigma-Aldrich, Missouri, USA), Docetaxel (Selleck Chemicals, Houston, USA), Cisplatin (Hospira UK Ltd).

### Sequential FACS (Fluorescent activated cell sorting) subpopulation isolation

Bulk HMLER and HCC38 cell lines were expanded *in vitro* in 150 mm tissue culture-treated culture dish (Corning plates). For the initial FACS rounds, cells were trypsinized and 1-2 × 10^7^ cells were stained for the membrane marker CD44 (BD anti-human CD44-PE-cy7 (1:800)) for 25 min at 4C. Antibody titration was previously determined staining a battery of breast cancer cell lines. Pure CD44^Hi^ and CD44^Lo^ cells were collected and replated for expansion in culture. Following sorting purification, cultures were supplemented with 0.1% (v/v) gentamicin and 1% (v/v) antibiotic-antimycotic for 2 passages to avoid contamination. Sequential rounds of FACS enrichment were performed until 100% pure populations were isolated (exemplified in schematic below). Purity was additionally confirmed functionally, as previously described ^14^. Data collection was performed using a BD Aria III and FACSDiva software (BD Biosciences). Flowjo X10.7.1 was used for data analysis

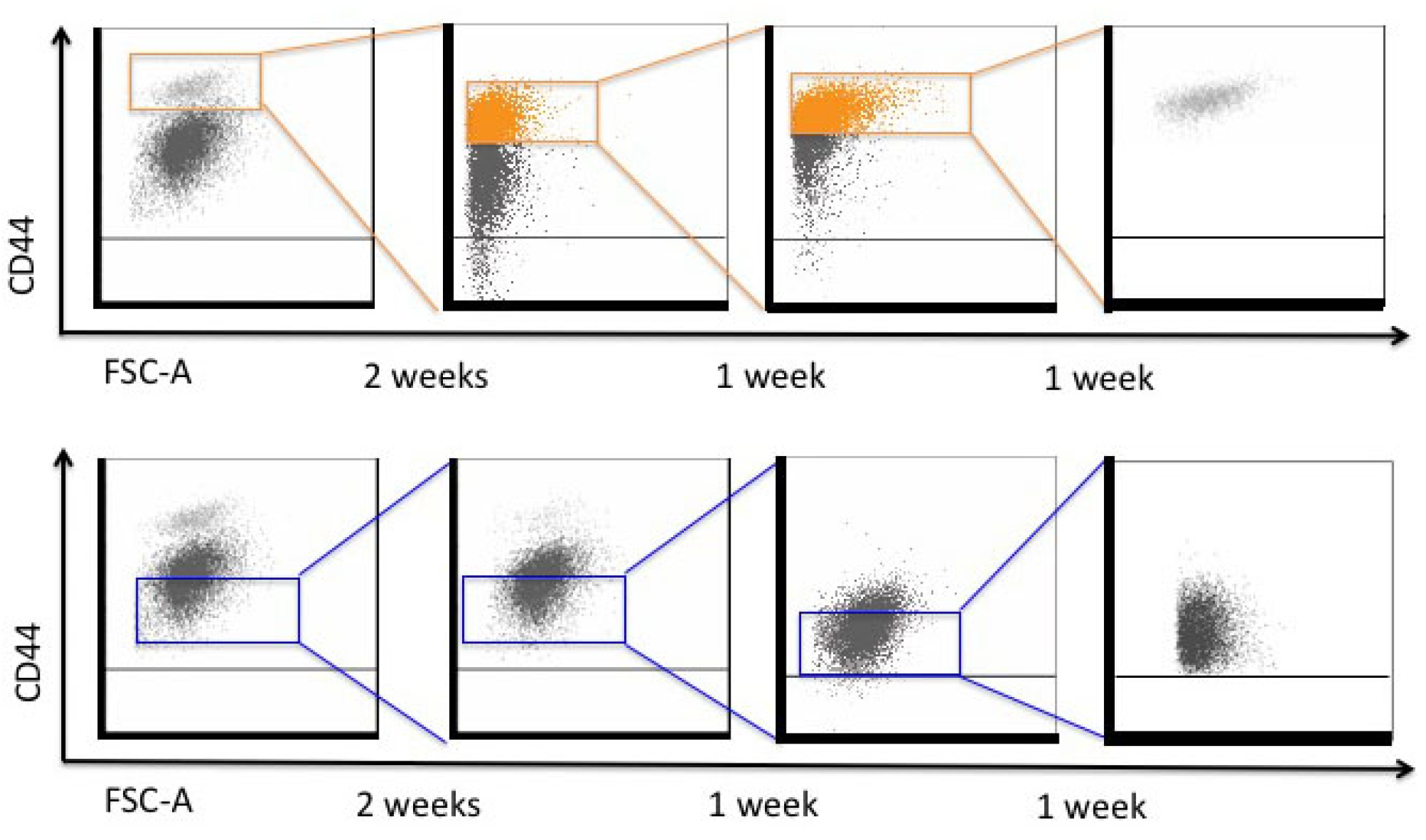

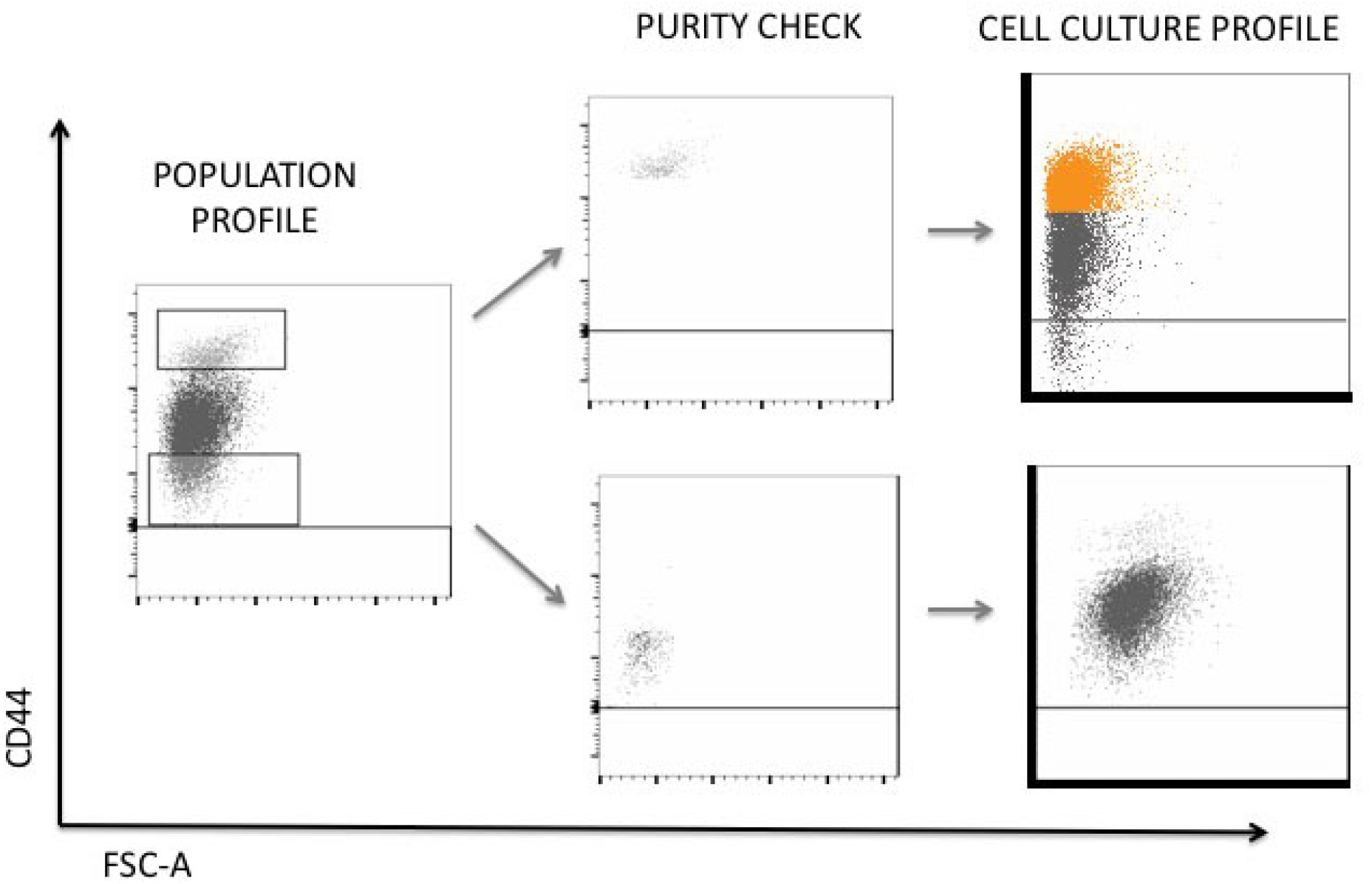

### Cell subpopulation RNA-seq analysis

Cells were grown in 150 mm tissue culture-treated culture dishes for 48h-72 h, depending on growth kinetics, till cells reached 80% confluence. At harvest time, cells were washed twice with cold PBS (Invitrogen), scraped and collected in 1.5 ml Eppendorf tubes. Cells were spun down, washed with cold PBS, and cell pellets snap freeze in liquid nitrogen. Frozen cell pellets (1 × 10^6^ cells) were used for RNA extraction using the commercial RNeasy Kit (Qiagen). Quality of RNA was assessed using a Bioanalyzer Eukaryote Total RNA Nano chip (Agilent). Samples that scored above RIN:9 were used to generate the mRNA library, using a TruSeq® Stranded mRNA LT - Set A kit (Illumina: RS-122-2101).

Three independent biological replicates of MCF-7, ZR75-1 and CD44^Lo^ versus CD44^Hi^ HMLER and HCC38 subpopulations were submitted for pair-end sequencing (Illumina HiSeq 2500 v4.0) at the Garvan Molecular Genetics Facility (Sydney, Australia). Raw files, read counts and normalised expression data can be downloaded from Gene Expression Omnibus (GEO), http://www.ncbi.nlm.nih.gov/geo, under accession GSE184647.

### Statistical Analysis of RNA-seq data

Paired-End 125 bp reads were aligned to the GRCh38 RefSeq build of the Homo sapiens (human) genome using the Subread aligner^49^. Genewise counts were obtained using featureCounts^50^. Genes were filtered from downstream analysis if they failed to achieve a CPM (counts per million mapped reads) value of at least 0.5 in at least three libraries. Counts were converted to log2-CPM, TMM normalized^51^ and precision weighted with the voom function of the limma package ^52, 53^. A linear model was fitted to each gene and a P value was computed against the null hypothesis for each gene using empirical Bayes moderated t-statistics^54^. P values were adjusted for multiple testing using the Benjamini-Hochberg method. A Differential Expression call was defined as a gene with a false discovery rate (FDR) below 5%.

### Ensemble Analysis

In addition to differential expression, we performed several analyses to identify genes involved in regulation of CD44^Lo^ to CD44^Hi^ plasticity.

#### Canonical Correlation Analysis (CCA)

Canonical Correlation Analysis^55^ involves finding a lower-dimensional representation of two datasets X and Y, where the correlation between X and Y is maximized. Let n represent the number of genes, p number of samples in X and q the number of genesets in Y. Assume that the columns of Xn×p and Ynxq have been centered and scaled. CCA finds up×r and vq×r, r < min(p,q), that maximize cor(Xu,Yv) - that is, it solves

Maximize u,vuTXTYv subject to uTXTXu ≤ 1, vTXTXv ≤1.

In our analyses, we asked what makes the CD44^Hi^ states in the HMLER and HCC338 cell lines correlated. We used the penalized CCA approach of ^56^ to quantify the contribution of genes to the correlation between the two datasets. CCA was applied to log-CPM values and a three-dimensional space of correlations was learned. However, only the genes with non-zero coefficients in the first canonical variates (CCA dimension) were considered as the genes contributing to the correlation between the two cell lines.

#### Nearest shrunken centroid analysis

We trained a Nearest shrunken centroid ^57^ classifier to identify genes that discriminate CD44^Hi^ and CD44^Lo^ states in the HMLER and HCC38 cell lines. The classifier was applied to log-CPM values. Optimal shrinkage threshold was determined by 5-fold cross-validation. Genes with non-zero scores were considered as genes contributing to differences between the CD44^Lo^ to CD44^Hi^ states.

The genes resulting from the above analyses were identified as a transcription factor if they were associated with DNA-binding transcription factor activity in Gene Ontology database (GO:0003700). Draggability was determined based on records in the DrugBank database ^58^. The results from these analyses were collectively used to rank genes displayed in the sankey plot. That is, every gene gets a vote if it is identified by any of the above analyses and criteria. Genes are then ranked by the majority vote.

### TCGA-CD44 cell-line mapping via Optimal Transport

Raw RNA sequencing count data for TCGA BRCA tumours and clinical annotation were downloaded using TCGA Biolinks Bioconductor package ^59^. RNA-seq counts were filtered by expression (using filterByExpr) and TMM normalised ^51^ using edgeR ^60^. Principal Component Analysis (PCA) was applied to log-counts (with +1 offset) in TCGA and cell-line data. We transported the 2nd to the fifth PCs (i.e. PC2-PC5) of the cell line data to PC3-PC6 of the TCGA data using Earth Mover Distance with Laplacian regularization^61^, as the two datasets exhibited similar topology on these dimensions, that is the relative distance of samples belonging to a subtype was similar on the selected PC dimensions. For example, the relative distance of the Basal samples to the Luminal samples is similar on the mentioned dimensions in both TCGA and cell line data. Given that Principal Components are constructed by linear combinations of genes, we related this topological similarity to transcriptional similarity. This approach of mapping datasets based on topological similarity is particularly helpful in small sample size scenarios such as our cell line data, as development of transcriptional signatures by machine learning models for assessment of similarity requires substantial data (samples) for training. Once cell line samples are mapped to TCGA samples, basal TCGA-BRCA samples similar to basal cell line samples were determined by a k Nearest Neighbour, k-NN, classifier. Here, the k was set to 5 - that is 5 nearest Basal TCGA samples to each of the basal cell line samples were considered as having a transcriptional profile similar to basal samples in the cell line data.

### Differential expression in TCGA breast cancer cohort

Raw RNA counts were obtained and processed as described in Statistical Analysis of RNA-seq data. Differential expression between inferred CD44^Lo^-like and CD44^Hi^-like samples was determined by limma-voom [4] at 5% FDR. We used limma to generate barcode plot and test for pathway enrichment.

### Survival Analysis

Kaplan Meier plot and survival analysis was done using survival ^62^ and survminer ^63^ R packages based on 151 TCGA BRCA basal subtype samples with valid survival data.

### The AR signalling gene regulatory network

Gene expression data of CD44^Hi^ and CD44_Lo_ mapped TCGA patients was used to build gene regulatory networks. The z-score method implemented in the dcanr package ^26^ was used to estimate the differential co-expression network between patients with a CD44^Hi^-like phenotype and patients with a CD44^Lo^-like phenotype at an FDR of 0.1. Interactions assessed were restricted to those involving at least 1 known transcription factor (TF) or TF co-regulator defined using the AnimalTFDB3.0 ^64.^ A CD44^Hi^-specific network was obtained by selecting those edges where the correlation in CD44^Lo^ samples was between −0.1 and 0.1. The network of CD44^Hi^ regulators was generated by inducing those edges from the CD44^Hi^-specific network that were between TF and their co-regulators and were evidenced by a protein-protein interaction. The protein-protein interaction network used to derive these networks was downloaded from the international molecular exchange (IMEx) ^65^. The CD44^Lo^-specific network and its associated regulator network was derived likewise.

### Immunofluorescence analysis

#### AR and ZEB1 expression analysis in cell subpopulations and cell lines

HMLER CD44^Lo^ cells (3 × 10^4^), HCC38 CD44^Lo^ (4 × 10^4^), MDA-MB-231 (1 × 10^4^), SUM159PT (1 × 10^4^) and HMLER/HCC38 CD44^Hi^ (1 × 10^4^) were seeded on 8 well chambers (Corning 8 well chamber 354118#) and cultured for 72 h. Cells were washed and fixed at the end-point.

#### AR and ZEB1 expression analysis in CD44^Lo^ cells in response to chemotherapy and AR antagonists

HMLER CD44^Lo^ cells (3 × 10^4^) were seeded on 8 well chambers (Corning 8 well chamber 354118#) and cultured for 72 h. Cells were treated 24 h post-seeding (0.1% (v/v) DMSO for control wells and 50 - 100 nM Dox) to assess chemotherapy effect on AR/ ZEB1 expression after 48 h drug exposure.

HCC38 CD44^Lo^ cells (3 – 6 × 10^4^) were seeded in 96 well plates. 24 h post-seeding cells were treated with chemotherapy (50 - 100 nM Dox, 1 - 2 μg/ml Cis, 1 – 4 nM Dtx), 10 μM AR antagonists (Sev, Abi or Enz) or combination treatments (50 nM Dox + 10 μM Sev, Abi or Enz; 2 μg/ml Cis + 10 μM Sev, Abi or Enz and 4 nM Dtx + 10 μM Sev, Abi or Enz) and cells were fixed for immunofluorescence analysis 48h post adding treatment.

HCC38 CD44^Hi^ cells (1 × 10^4^) were seeded in 96 well plates. 24 h post-seeding cells were treated with 10 μM AR antagonists (Sev, Abi or Enz). Cells were fixed 48h post treatment.

### Immunofluorescence staining

Cells were fixed with 4% paraformaldehyde (Electron Microscopy Sciences) for 15 min and permeabilized with 0.3% (v/v) Triton X-100 for 20 min at RT. Following fixation, cells were blocked with 3% (w/v) bovine serum albumin (BSA) and 5% (v/v) Horse Serum in PBS and stained with the primary antibodies: mouse anti-AR441 (DAKO, 1:200), rabbit anti-ZEB1 (Santa Cruz #H-102, 1:200) for 1 h at room temperature or overnight at 4oC. Cell were washed three times with 0.2% (v/v) Tween-20, followed by incubation with the appropriate secondary antibodies conjugated with Cyanine Cy3, Alexa Fluor 647, or with Phalloidin-iFluor 488 or 647, and processed as described. Coverslips/chambers were mounted with ProLong Diamond Antifade Mounting Media (ThermoFisher Scientific). Alternatively, cells grown in 96-well glass-bottomed plates were imaged in 80% glycerol post-labelling. Fluorescence was recorded via confocal laser-scanning microscopy using either a Leica DMI 6000 SP8 with 40x (NA 1.3) or 63x (NA 1.4) oil objectives or a Nikon A1R confocal with 20x Plan Apochromat air objective (NA 0.75) at 2x zoom using an HD25 resonance scanner. All images (Lift, Leica; ND2, Nikon) were converted to TIFF format prior to quantitative analysis. Brightness and contrast were optimized with Fiji software (National Institutes of Health) for visualisation / presentation only.

### Immunofluorescence Image Analysis

Image quantification was performed using CellProfiler Analyst software ^66^ (v4.0 or higher; www.cellprofiler.org). Cells were individually segmented using DAPI and Phalloidin as nuclear and cell body markers, respectively. In the absence of Phalloidin staining, AR and ZEB1 channel data was summed to create a 4th channel enabling improved detection of complete cell bodies.

### Image-derived Quantitative Data Analysis

Segmented images and single cell quantitative data were integrated, parsed and analysed using custom-developed Knime workflows^67^ (KNIME AG, Zurich). Quantitative cell data was grouped by individual wells and outlier cells were detected and removed by means of interquartile range, R = [Q_1_ - k(IQR), Q_3_ + k(IQR)] with IQR = Q_3_ - Q_1_ and k = 4, using AR and ZEB1 integrated cell body intensities. All quantitative feature distributions were then normalized via Z-score normalization. Publication plots were generated in Jupyter notebook^68^ using the Python packages ‘matplotlib’^69^, ‘numpy’^70^, ‘scipy’ ^71^ and ‘seaborn’^72^.

### Tumorsphere assay

Single cell suspensions were plated in ultra-low attachment 96-well plates (Corning # CLS3474, New York, USA) at low densities optimized to ensure tumorspheres arose from single anchor-independent cells. Cells were seeded in 100μl at the following density: HMLER-CD44^Hi^ (50 cells/well), HCC38 CD44^Hi^, MDA-MB-231 and SUM159PT (100 - 200 cells/well). Cell line specific serum-free media was supplemented with 1% (v/v) penicillin/streptomycin, 20 ng/ml EGF, 20 ng/ml FGFb, 4 μg/mL heparin, 1x B27 and 1% (v/v) methyl cellulose (Sigma-Aldrich). Fresh media was topped up every 5 days by adding 50 μl per well of the appropriate tumorsphere media. Tumorspheres were counted directly or pictures were taken for digital/manual quantitation and size analysis. Tumorspheres were analysed at day 7 (HCC38 siRNA assay), day 10 (HMLER and SUM159PT) and day 14 and 21 (HCC38, MDA-231), respectively. Two to four independent assays were carried out per cell line/assay, 10-15 independent wells per condition and assay were counted. Results are presented as tumorsphere formation efficiency (number of spheres counted divided by the number of cells seeded per well).

### Western Blot analysis

Protein was extracted from cell lysates using RIPA lysis buffer (San Cruz, sc-24948) or specific lysis buffer for phospho-protein detection containing 20mM Tris-HCl ph7.6, 137 mM NaCl, 1% (v/v) NP40, 0.5% (v/v) Na-deoxycholate, 10 mM NaF, 20 mM b-glycerophosphate, 1 mM Na-orthovanadate and 1:100 (v/v) Halt protease inhibitor cocktail (ThermoFisher). 30 μg of protein were loaded and run in 4-12% Bis-tris gels (Invitrogen). Proteins were transferred to a PVDF membrane (BioRad). After blocking with 5% (w/v) BSA and 0.05% (v/v) Tween-20 in PBS, the membranes were incubated overnight with primary antibodies diluted in 5% (w/v) BSA. The following antibodies were used in this study: Rabbit anti-E-Cadherin (24E10) mAb #3195 (cell signalling 1:1000), rabbit anti-ZEB1 (Santa Cruz #H-102, 1:1000), mouse anti-AR 441 (DAKO, 1:1000), mouse anti-Tubulin (SIGMA-Aldrich, #T6199 clone DM1A, 1:2000) and mouse anti-GAPDH (CST D4C6R#97166). Following antibody incubation membranes were washed with 0.05% (v/v) Tween-20 in PBS and incubated with the corresponding horseradish peroxidase-conjugated secondary antibodies (CST, 1/5000) for 30 min. Blots were developed using Western Lightning Plus ECL (PerkinElmer) and FusionFx7 Digital Imager. Band intensities were quantified using Fiji and results were represented relative to controls. At least two independent biological assays were used for the analysis.

### Drug response assay

CD44^Hi^ HMLER, HCC38 and MDA-MB-231 cells were plated in triplicate in 96-well plates (500 cells per well). After 24 hours, cells were treated with control (0.1% DMSO (v/v)) or AR antagonist: seviteronel, enzalutamide or abiraterone (0.01, 0.6, 1.2, 2.5, 5, 10, 20, 40, 80 and 160 μM). Three independent assays were run. Cell viability was determined using the alamarBlue assays for cell viability assay according to manufacturer’s protocol (ThermoFisher Scientific). Absorbance was measured at 470nm using an EnSipre Multimode plate reader (Perkim Elmer). Percentage viability was determined as per reference to untreated cells.

### TNBC AR targeted gene set analysis RNA-seq

HCC38 CD44^Hi^ cells (3.5 × 10^5^) were plated in P10s, 24h post seeding cells were treated at with siCtrl (30 nM), siAR (30 nM (1:1) Ex1/Ex7), vehicle (0.1% DMSO (v/v)) or 10 μM seviteronel, abiraterone and enzalutamide. siRNAs were transfected using Optimem Reduced Serum Media-(ThermoFisher 31985070) and Lipofectamine 3000 (ThermoFisher scientific L3000015#) as per standard protocol. 48h post-treatment cells were harvested and RNA extracted as described in RNA-seq section above. Three independent biological samples were submitted for pair-end sequencing (Illumina NovaSeq 6000 System) at Macrogen Oceania PL. Data can be retrieved from GEO, under accession GSE184455.

### Data representation

For visually combining gene expression with functional annotation, we used the packages GOplot^73^ and clusterProfiler^74^ in R. AR targets were tested on terms from the Molecular Signatures Database (MSigDB), Broad Institute (http://software.broadinstitute.org), matching the top 1000 differentially expressed genes from the comparison between siAR versus siCtrl and Sev, Abi or Enz versus control (DMSO). Pathway enrichment analysis across treated samples, we used the Reactome Knowlegbase and KEGG Pathways database.

### Chemotherapy-induced plasticity assays (Flow Cytometry)

#### HMLER cell line

##### 72h assay

2 × 10^5^ pure HMLER CD44^Lo^ cells were seeded in 60mm cell culture plates. Cells were treated 24h post seeding with low and high dose chemotherapy (50 - 100 nM Dox, 1 - 2 μg/ml Cis, 1 – 4 nM Dtx); 10 μM AR antagonists (Sev or Enz) or combination treatments (50 nM Dox + 10 μM Sev; 2 μg/ml Cis + 10 μM Sev and 4 nM Dtx + 10 μM Sev). 72h following treatment addition, cells were trypsinized and stained to define expression levels of the membrane marker CD44. Cells were stained in FACS buffer (2% (v/v) FBS in PBS) and anti-human CD44 PE-Cy7 (1:800) for 25 min at 4°C. Cell viability was determine using the Zombie Aqua™ Fixable Viability Kit (Biolegend) as per reference protocol. After primary antibody and viability labelling, cells were washed in FACS buffer twice, fixed in 4% (v/v) paraformaldehyde for 15 min, washed twice in FACS buffer and stored at 4°C up to 1 week. Data collection was performed using a BD LSR Fortessa. Flowjo X10.7.1 was used for data analysis and Graphpad Prism 9 for graphing and statistical analysis. Three independent biological assays were performed and analyzed.

##### 48h-120h assay

8.5 × 10^5^ pure HMLER CD44^Lo^ cells were seeded in 100mm cell culture plates. 24h post seeding cells were treated with 0.1% (v/v) DMSO for controls, low and high chemotherapy doses (25 - 50 nM Dox, 1 - 2 μg/ml Cis, 1 – 4 nM Dtx), 10 μM AR antagonists (Sev or Enz) or combination treatments (25 - 50 nM Dox + 10 μM Sev or Enz; 1 - 2 μg/ml Cis + 10 μM Sev or Enz and 1 - 4 nM Dtx + 10 μM Sev or Enz). 48h post treatment cells were trypsinized and 1 × 10^5^ cells were analysed by flow cytometry to determine CD44 levels. From the remaining cells, 4.5 × 10^5^ cells were re-seeded and allowed to recover for 24h. Treatment was added for additional 72h (120h total treatment). Cells were trypsinized at that time point and, when enough cells were available, 1 × 10^5^ cells were stained for subsequent flow cytometry analysis. Analysis of CD44 levels and viability was performed as described above. Two independent biological assays were performed and analyzed.

#### HCC38 cell line

##### 72h assay

1 × 10^6^ pure HCC38 CD44^Lo^ cells were seeded in 60mm cell culture plates. Cells were treated 24h post seeding with (50 - 100 nM Dox) or combination treatments (50 nM Dox + 10 μM Sev, Abi or Enz). 72h following treatment addition, cells were trypsinized and stained for the membrane marker CD44 as described above. Three independent biological assays were performed and analyzed.

##### 144h assay

4.5 × 10^5^ pure HCC38 CD44^Lo^ cells were seeded 60mm cell culture plates. Cells were treated 24h post seeding with (50 - 100 nM Dox, 1 - 2 μg/ml Cis, 1 – 4 nM Dtx), 10 μM AR antagonists (Sev, Abi and Enz) or combination treatments (50 nM Dox + 10 μM Sev, Abi or Enz; 2 μg/ml Cis + 10 μM Sev, Abi or Enz and 4 nM Dtx + 10 μM Sev, Abi or Enz). 72h following treatment addition, cells were trypsinized and stained for the membrane marker CD44 as described above. Four independent biological assays were analyzed.

The gating strategy followed is exemplified below. The panel shows the profiles of untreated CD44^Lo^ HCC38 cells (top) versus Dox treated cells (bottom) at experimental end-point. The plots on the far-right side show spontaneous (top panel) versus chemotherapy-induced (bottom panel) de novo formed CD44^Hi^ cells. The top edge of the bulk CD44^Lo^ population from the untreated control was used to define the threshold between CD44^Lo^ and CD44^Hi^ cells for all conditions.

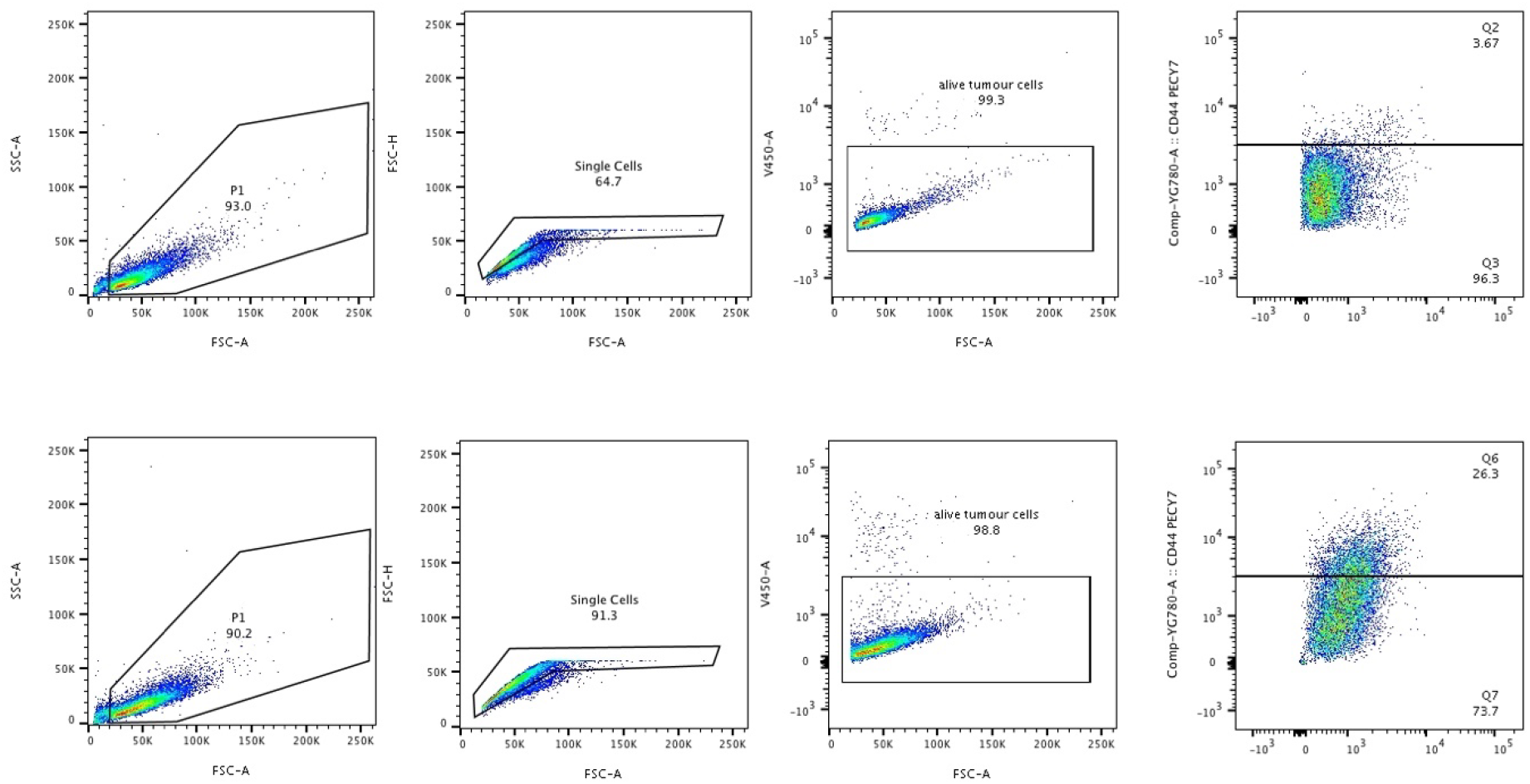

### Animal Studies

NOD/SCID mice at 6-8 weeks of age were purchased from Australian Bioresources (ABR). Mice were 8-9 weeks of age at time of injections. For orthotopic injections of cell lines, cells (1×10^6^ cells) were prepared in 25 μl 20% Matrigel (BD Matrigel Matrix Growth Factor Reduced (GFR) / serum-free media and injected into the inguinal mammary fat pads. For PDX specimens, freshly passaged single tumour chunks (1mm^3^) were implanted orthotopically. All experiments were approved by and conducted in accordance with the National Health and Medical Research Council Statement on Animal Experimentation, the requirements of new South Wales State Government legislation, and with the rules for animal experimentation of the Biological Testing Facility of the Garvan Institute and the Victor Chang Cardiac Research Institute (protocol #18/12).

Twelve to fifteen animals were enrolled per treatment arm for each experiment. Animals were randomized into treatment arms (specified for each experiment) at the time tumours ranged between 50-150mm^3^. Tumour volume was estimated by calliper measurements twice per week up until the harvest date. AR antagonists were administrated by oral gavage (100 mg/kg) daily for 1 month per treatment cycle, unless otherwise specified. Two weeks between cycles were allowed for animals to recover. AR antagonists were resuspended in 1% (w/v) carboxymethylcellulose, 0.1% (v/v) Tween 80, 5% (v/v) DMSO. Vehicle or chemotherapy single treatment arms were administrated diluent by daily oral gavage for the duration of the treatment cycles. Chemotherapy (20 mg/kg Dox, 4 mg/kg PEG-Dox, 10 mg/ml Cis, 20 mg/kg Ptx or 40 mg/kg NAB-Ptx) was administrated once per week via intraperitoneal injections 1 week following AR antagonists / diluent gavaging starting date. Vehicle and chemotherapy single treatment arms received saline injections. Animals in vehicle or single AR antagonist treatment arms were harvested at conclusion of cycle 1 of treatment or when reached ethical end-point (1000mm^3^ or tumour burden greater than 10% of animal body weight). For chemotherapy or combination chemotherapy plus AR antagonist treatment arms, half of the animals were harvested at end of either cycle 1, 2 and/or 3 to determine treatment effects on tumour growth and metastatic incidence. The remaining animals were used to track post-treatment tumour evolution as indicated for each experiment in the results section. Metastatic evolution was tracked in xenograft models expressing Luciferase using IVIS-spectrum imaging weekly or at harvest time. For weekly tracking, animals were shaved, weighed and subsequently injected with 10 μl/g of D-luciferin Potassium Salt stock (15 mg/ml, Sigma, LUCK-2G) subcutaneously. 3 minutes after D-luciferin administration animals were imaged (auto-exposure and 1sec. C and B magnification) to record metastatic burden in the presence and absence of primary tumour. To define early metastatic burden at harvest point, animals were weighted and injected luciferin as specified above. 10 minutes after D-luciferin administration, animals were sacrificed, removing the primary tumour to detect peritoneum and small lymph node metastasis. Afterwards the chest cavity was opened to expose internal organs for signal detection to determine overall metastatic incidence using total flux measurements.

### PDX models FACS profiling

Tumours were harvested when reached 800 mm3 volume, chopped into small pieces, then incubated at 37°C for 40 min on a rotary shaker in DMEM/F12 containing Collagenase A (300Units) and Hyaluronidase (100Units). Following digestion, tumour cell suspensions were pelleted, the DME removed, washed with PBS 2% (v/v) FBS and then resuspended in 0.15% + 10% DNAseI trypsin for 1 min. Trypsin was quenched with 2%FBS/DME. Cells were treated with 1 mg/ml DNAseI, then filtered through a 40 μm filter. Cells were stained and analysed by flow cytometry or stored in freezing medium (10% DMSO, 90% Calf serum). Cell pellets were stained for FACS analysis using a panel of antibodies previously established in the lab (CD298-PE bio-legend (1:200), CD44-PEcy7 BD-Bioscience (1:800)). Samples were analysed on a BD Bioscience Fortessa. Results were analysed and using Flowjo10.7.1. Gating strategy exemplified below.

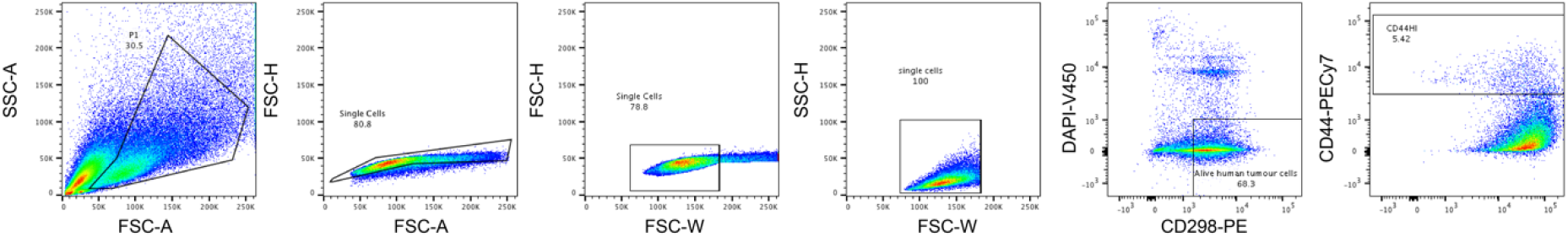

### Immunohistochemistry

Tissues were fixed in 10% Neutral Buffered Formalin for 24 hours, embedded in paraffin following standard protocol and processed on the Leica Peloris II. Slides (4 μm sections) were deparaffinised with xylene and rehydrated with washes in decreasing concentrations of ethanol, performed on the Leica Autostainer XL. The same autostainer was used for H&E staining and counterstaining with Haematoxylin (Shandon Instant Haematoxylin Kit). Stainings for Cleaved Caspase-3 and Ki67 were performed with a Leica Bond RX fully automated research stainer and the Leica Bond Polymer Refine Detection kit. Heat-induced epitope retrieval with Bond Epitope Retrieval Solution 2 (EDTA based, pH 9) at 93°C for 30 min and 20 min was performed for Ki67 and CC3 staining respectively and at 93°C for 30 minutes for ZEB1. The slides were stained according to the IHC-F 60 min Ab incubation protocol using either a 1:500 dilution of the Ki67 Rabbit antibody (ThermoScientific), a 1:200 dilution of the Cleaved Caspase-3 Polyclonal Rabbit antibody (Cell Signaling) or 1:200 for ZEB1 Polyclonal Rabbit antibody (Cell signalling (H-102)) as primary antibodies. IHC quantification for Ki67 and CASP3 was performed using QuPath imaging analysis software (v0.2.3). Tumour cells within tissue were selected for by object classification, trained using known regions of tumour cells. A watershed cell detection based on the hematoxylin counterstain was used to identify nuclei within these regions. Average DAB optical density was then computed for each cell and classed as either positive or negative for DAB staining using a consistent threshold. The average of 5 to 7 tumours per condition was used for quantification.

AR mouse monoclonal antibody (AR 441, DAKO M3562#) staining was performed using a manual DAKO auto-stainer. Antigen retrieval (S2367) was run for 10 sec, antibody was then incubated for 60 min (1:250) and coupled with the novolink secondary detection system. Initial optimization included a positive control (normal breast tissue) and a negative control (lung and kidney tissue) to ensure the specificity of the staining. Each tumour core was scored by a pathologist for percentage of positive tumour cells, for staining intensity (1+, 2+ 3+) and for localization (nuclear, cytoplasmic).

### Statistical analysis of immunohistochemical staining for AR and ZEB1 in clinical cohorts

#### Treatment-naïve TNBC clinical cohorts

Tissue Microarrays representing 308 treatment-naive triple-negative breast cancer specimens collected from the Royal Prince Alfred Hospital, Concord Repatriation General Hospital and Royal North Shore Hospital in New South Wales, Australia. The studies were approved by the Royal Prince Alfred Hospital Human Ethics Review Committee (X14-0241) and by the Northern Sydney Local Health District Human Research Ethics Committee (X15-0388). A multivariate analysis was done considering all prognostic indicators. Multiple measurements for each patient tumour (independent cores and independent scores) were collapsed to generate a maximum hybrid score for each patient (where Hybrid scores are staining intensity * percentage / 3, where score = (“percent 1+” + 2*”percent 2+” + 3*”percent 3+”)/3).

#### SETUP trial clinical cohort

Samples were stained and scored as above. The study was approved by the human research and ethics committees at all the participating institutions (Monash Medical Centre, Melbourne, Australia and Monash University, Melbourne, Australia) under protocol 03169A. All participating women provided written and informed consent

#### Staining of PDXs and patient samples

Ethical approval for AR and ZEB1 staining on human clinical cohorts and patient-derived xenograft samples was obtained and approved by the St Vincent’s Hospital Human Research Ethics Committee (2019/ETH12832).

#### Post-hoc biomarker analysis of Clarity-01 Phase II clinical trial

Data used for this analysis are from the Clarity-01 clinical trial (IND133101, NCT02580448). The study was approved by the institutional review board at each participating site. Informed consent was obtained from all individual participants included in the study.

Participating sites include:

1. Memorial Sloan Kettering Cancer Center
2. Gabrail Cancer Center
3. Duke University Hospital System
4. University of Colorado Cancer Center
5. Henry Ford Hospital
6. Massachusetts General Hospital
7. University of Texas Southwestern Medical Center
8. Virginia Oncology Associates
9. North Shore Hematology Oncology Associates, P.C.
10. The Ohio State University, Stefanie Spielman Comprehensive Breast Center
11. University of Louisville Hospital, James Graham Brown Cancer Center
12. Huntsman Cancer Institute
13. Hackensack University Medical Center
14. Oregon Health & Science University Community Hematology Oncology - Northwest
15. UNC Hospitals, The University of North Carolina at Chapel Hill
16. University of Alabama at Birmingham
17. University of Michigan
18. Mary Crowley Cancer Research Centers - Medical City
19. University of Minnesota Medical Center, Fairview
20. Charleston Hematology Oncology Associates, PA
21. Georgia Cancer Center at Augusta University
22. The Sarah Cannon Research Institute
23. Florida Cancer Specialists
24. Florida Cancer Specialists
25. US Oncology, Inc. / Texas Oncology - Baylor Charles A. Sammons Cancer Center
26. US Oncology, Inc. / Oncology Hematology Care, Inc.
27. US Oncology, Inc. / Nebraska Cancer Specialists
28. Precision Cancer Research / Brig Center for Cancer Care and Survivorship, LLC
29. US Oncology, Inc. / Rocky Mountain Cancer Centers
30. City of Hope (City of Hope National Medical Center, City of Hope Medical Center)

Cytoplasmic and nuclear AR expression were scored as percentage of positive cells at different staining intensities (1+, 2+, 3+) converted to H-scores as 1 × (% cells at 1+) + 2 × (% cells at 2+) + 3 × (% cells at 3+). Patients were split into two groups with low and high expression, using H-score cut-offs as indicated. Kaplan-Meier curves were produced for each group, and median survival times derived. Hazard ratios and 95% confidence intervals were estimated using a univariable Cox proportional-hazards model with binarized expression as explanatory variable.

## Notes

### Funding Statement

This study was funded by a St. Vincents Hospital Research Grant, NHMRC Ideas grants APP1088122 and APP1181230, a National Breast Cancer Foundation research grant IIRS-19-092. L.R. is supported by a Ramon-Areces Fellowship. C.L.C and B.P.S.J were supported by the Nelune Foundation Rebecca Wilson Fellowship. C.L.C was supported by a CINSW Fellowship CDF181243. L.D.G. was supported by the Kinghorn Foundation. TEH was supported by a National Breast Cancer Foundation fellowship (IIRS-19-009). FVK and CAP are supported by an Australian Government Research Training Program (RTP) Scholarship.

### Author Declarations

Animal Experiments All experiments were approved by and conducted in accordance with the National Health and Medical Research Council Statement on Animal Experimentation, the requirements of New South Wales State Government legislation, and with the rules for animal experimentation of the Biological Testing Facility of the Garvan Institute and the Victor Chang Cardiac Research Institute (protocol #18/12). Treatment-naive TNBC clinical cohorts Tissue Microarrays representing 308 treatment-naive triple-negative breast cancer specimens collected from the Royal Prince Alfred Hospital, Concord Repatriation General Hospital and Royal North Shore Hospital in New South Wales, Australia. The studies were approved by the Royal Prince Alfred Hospital Human Ethics Review Committee (X14-0241) and by the Northern Sydney Local Health District Human Research Ethics Committee (X15-0388). SETUP trial clinical cohort Samples were stained and scored as above. The study was approved by the human research and ethics committees at all the participating institutions (Monash Medical Centre, Melbourne, Australia and Monash University, Melbourne, Australia) under protocol 03169A. Staining of PDXs and patient samples Ethical approval for AR and ZEB1 staining on human clinical cohorts and patient-derived xenograft samples was obtained and approved by the St Vincent's Hospital Human Research Ethics Committee (2019/ETH12832). Post-hoc biomarker analysis of Clarity-01 Phase II clinical trial Data used for this analysis are from the Clarity-01 clinical trial (IND133101, NCT02580448). The study was approved by the institutional review board at each participating site. Informed consent was obtained from all individual participants included in the study. Participating sites include: 1.Memorial Sloan Kettering Cancer Center 2.Gabrail Cancer Center 3.Duke University Hospital System 4.University of Colorado Cancer Center 5.Henry Ford Hospital 6.Massachusetts General Hospital 7.University of Texas Southwestern Medical Center 8.Virginia Oncology Associates 9.North Shore Hematology Oncology Associates, P.C. 10.The Ohio State University, Stefanie Spielman Comprehensive Breast Center 11.University of Louisville Hospital, James Graham Brown Cancer Center 12.Huntsman Cancer Institute 13.Hackensack University Medical Center 14.Oregon Health & Science University Community Hematology Oncology - Northwest 15.UNC Hospitals, The University of North Carolina at Chapel Hill 16.University of Alabama at Birmingham 17.University of Michigan 18.Mary Crowley Cancer Research Centers - Medical City 19.University of Minnesota Medical Center, Fairview 20.Charleston Hematology Oncology Associates, PA 21.Georgia Cancer Center at Augusta University 22.The Sarah Cannon Research Institute 23.Florida Cancer Specialists 24.Florida Cancer Specialists 25.US Oncology, Inc. / Texas Oncology - Baylor Charles A. Sammons Cancer Center 26.US Oncology, Inc. / Oncology Hematology Care, Inc. 27.US Oncology, Inc. / Nebraska Cancer Specialists 28.Precision Cancer Research / Brig Center for Cancer Care and Survivorship, LLC 29.US Oncology, Inc. / Rocky Mountain Cancer Centers 30.City of Hope (City of Hope National Medical Center, City of Hope Medical Center)

## References

1. Hanahan, D. Hallmarks of Cancer: New Dimensions. Cancer discovery 12, 31 (2022).

2. Castano, Z. et al. IL-1beta inflammatory response driven by primary breast cancer prevents metastasis-initiating cell colonization. Nat Cell Biol 20, 1084–1097 (2018).

3. Landsberg, J. et al. Melanomas resist T-cell therapy through inflammation-induced reversible dedifferentiation. Nature 490, 412–416 (2012).

4. Malladi, S. et al. Metastatic Latency and Immune Evasion through Autocrine Inhibition of WNT. Cell 165, 45–60 (2016).

5. Ouzounova, M. et al. Monocytic and granulocytic myeloid derived suppressor cells differentially regulate spatiotemporal tumour plasticity during metastatic cascade. Nature communications 8, 14979 (2017).

6. Su, S. et al. A positive feedback loop between mesenchymal-like cancer cells and macrophages is essential to breast cancer metastasis. Cancer cell 25, 605–620 (2014).

7. Rambow, F. et al. Toward Minimal Residual Disease-Directed Therapy in Melanoma. Cell 174, 843-855.e819 (2018).

8. Su, Y. et al. Single-cell analysis resolves the cell state transition and signaling dynamics associated with melanoma drug-induced resistance. Proceedings of the National Academy of Sciences 114, 13679–13684 (2017).

9. Lambert, A.W. & Weinberg, R.A. Linking EMT programmes to normal and neoplastic epithelial stem cells. Nat Rev Cancer 21, 325–338 (2021).

10. Marine, J.C., Dawson, S.J. & Dawson, M.A. Non-genetic mechanisms of therapeutic resistance in cancer. Nat Rev Cancer (2020).

11. Sottoriva, A. et al. A Big Bang model of human colorectal tumor growth. Nature genetics 47, 209–216 (2015).

12. Esposito, M., Ganesan, S. & Kang, Y. Emerging strategies for treating metastasis. Nature cancer 2, 258–270 (2021).

13. Chaffer, C.L. et al. Normal and neoplastic nonstem cells can spontaneously convert to a stem-like state. Proc Natl Acad Sci U S A 108, 7950–7955 (2011).

14. Chaffer, C.L. et al. Poised chromatin at the ZEB1 promoter enables breast cancer cell plasticity and enhances tumorigenicity. Cell 154, 61–74 (2013).

15. Nieto, M.A., Huang, R.Y., Jackson, R.A. & Thiery, J.P. EMT: 2016. Cell 166, 21–45 (2016).

16. Goldman, A. et al. Temporally sequenced anticancer drugs overcome adaptive resistance by targeting a vulnerable chemotherapy-induced phenotypic transition. Nature communications 6, 6139 (2015).

17. Redfern, A.D., Spalding, L.J. & Thompson, E.W. The Kraken Wakes: induced EMT as a driver of tumour aggression and poor outcome. Clin Exp Metastasis 35, 285–308 (2018).

18. Tam, W.L. & Weinberg, R.A. The epigenetics of epithelial-mesenchymal plasticity in cancer. Nature medicine 19, 1438–1449 (2013).

19. Hu, X. et al. Induction of cancer cell stemness by chemotherapy. Cell cycle (Georgetown, Tex.) 11, 2691–2698 (2012).

20. Carey, L.A. et al. The triple negative paradox: primary tumor chemosensitivity of breast cancer subtypes. Clinical cancer research : an official journal of the American Association for Cancer Research 13, 2329–2334 (2007).

21. Liedtke, C. et al. Response to neoadjuvant therapy and long-term survival in patients with triple-negative breast cancer. Journal of clinical oncology : official journal of the American Society of Clinical Oncology 26, 1275–1281 (2008).

22. Deluche, E. et al. Contemporary outcomes of metastatic breast cancer among 22,000 women from the multicentre ESME cohort 2008-2016. European journal of cancer 129, 60–70 (2020).

23. Pastushenko, I. et al. Identification of the tumour transition states occurring during EMT. Nature 556, 463–468 (2018).

24. Aiello, N.M. et al. EMT Subtype Influences Epithelial Plasticity and Mode of Cell Migration. Developmental cell 45, 681-695.e684 (2018).

25. CGAN Comprehensive molecular portraits of human breast tumours. Nature 490, 61–70 (2012).

26. Bhuva, D.D., Cursons, J., Smyth, G.K. & Davis, M.J. Differential co-expression-based detection of conditional relationships in transcriptional data: comparative analysis and application to breast cancer. Genome Biol 20, 236 (2019).

27. Bleach, R. & McIlroy, M. The Divergent Function of Androgen Receptor in Breast Cancer; Analysis of Steroid Mediators and Tumor Intracrinology. Frontiers in endocrinology 9, 594 (2018).

28. Hickey, T.E., Robinson, J.L., Carroll, J.S. & Tilley, W.D. Minireview: The androgen receptor in breast tissues: growth inhibitor, tumor suppressor, oncogene? Molecular endocrinology (Baltimore, Md.) 26, 1252–1267 (2012).

29. Hickey, T.E. et al. The androgen receptor is a tumor suppressor in estrogen receptor-positive breast cancer. Nature medicine (2021).

30. Traina, T.A. et al. Enzalutamide for the Treatment of Androgen Receptor-Expressing Triple-Negative Breast Cancer. Journal of clinical oncology : official journal of the American Society of Clinical Oncology 36, 884–890 (2018).

31. Gucalp, A. et al. Phase II trial of bicalutamide in patients with androgen receptor-positive, estrogen receptor-negative metastatic Breast Cancer. Clinical cancer research : an official journal of the American Association for Cancer Research 19, 5505–5512 (2013).

32. Scher, H.I. et al. Increased Survival with Enzalutamide in Prostate Cancer after Chemotherapy. New England Journal of Medicine 367, 1187–1197 (2012).

33. de Bono, J.S. et al. Abiraterone and Increased Survival in Metastatic Prostate Cancer. New England Journal of Medicine 364, 1995–2005 (2011).

34. Bardia, A. et al. Phase 1 study of seviteronel, a selective CYP17 lyase and androgen receptor inhibitor, in women with estrogen receptor-positive or triple-negative breast cancer. Breast cancer research and treatment 171, 111–120 (2018).

35. Dontu, G. et al. In vitro propagation and transcriptional profiling of human mammary stem/progenitor cells. Genes & development 17, 1253–1270 (2003).

36. Davies, A. et al. An androgen receptor switch underlies lineage infidelity in treatment-resistant prostate cancer. Nat Cell Biol 23, 1023–1034 (2021).

37. Ishibashi, H. et al. Sex steroid hormone receptors in human thymoma. J Clin Endocrinol Metab 88, 2309–2317 (2003).

38. Ricciardelli, C. et al. The Magnitude of Androgen Receptor Positivity in Breast Cancer Is Critical for Reliable Prediction of Disease Outcome. Clinical cancer research : an official journal of the American Association for Cancer Research 24, 2328–2341 (2018).

39. Zaborowski, M., Pearson, A., Sioson, L., Gill, A.J. & Ahadi, M.S. Androgen receptor immunoexpression in triple-negative breast cancers: is it a prognostic factor? Pathology 51, 327–329 (2019).

40. Alamgeer, M. et al. Changes in aldehyde dehydrogenase-1 expression during neoadjuvant chemotherapy predict outcome in locally advanced breast cancer. Breast Cancer Research 16, R44 (2014).

41. Gucalp, A. et al. Phase (Ph) 2 stage 1 clinical activity of seviteronel, a selective CYP17-lyase and androgen receptor (AR) inhibitor, in women with advanced AR+ triple-negative breast cancer (TNBC) or estrogen receptor (ER)+ BC: CLARITY-01. Journal of Clinical Oncology 35, 1102–1102 (2017).

42. Robson, M. et al. Olaparib for Metastatic Breast Cancer in Patients with a Germline BRCA Mutation. The New England journal of medicine 377, 523–533 (2017).

43. Bardia, A. et al. Sacituzumab Govitecan in Metastatic Triple-Negative Breast Cancer. The New England journal of medicine 384, 1529–1541 (2021).

44. Bonnefoi, H. et al. A phase II trial of abiraterone acetate plus prednisone in patients with triple-negative androgen receptor positive locally advanced or metastatic breast cancer (UCBG 12-1). Annals of oncology : official journal of the European Society for Medical Oncology 27, 812–818 (2016).

45. Chia, K. et al. Non-canonical AR activity facilitates endocrine resistance in breast cancer. Endocr Relat Cancer 26, 251–264 (2019).

46. Norris, J.D. et al. Androgen receptor antagonism drives cytochrome P450 17A1 inhibitor efficacy in prostate cancer. The Journal of clinical investigation 127, 2326–2338 (2017).

47. Ellison, S.J. et al. Abstract P3-14-04: Effects of the dual selective CYP17 lyase inhibitor and androgen receptor (AR) antagonist, VT-464, on AR+ and ER+ tumor models in vitro and in vivo. Cancer research 76, P3-14-04 (2016).

48. Piccart-Gebhart, M.J. et al. Trastuzumab after Adjuvant Chemotherapy in HER2-Positive Breast Cancer. New England Journal of Medicine 353, 1659–1672 (2005).

49. Liao, Y., Smyth, G.K. & Shi, W. The Subread aligner: fast, accurate and scalable read mapping by seed-and-vote. Nucleic Acids Res 41, e108 (2013).

50. Liao, Y., Smyth, G.K. & Shi, W. featureCounts: an efficient general purpose program for assigning sequence reads to genomic features. Bioinformatics (Oxford, England) 30, 923–930 (2014).

51. Robinson, M.D. & Oshlack, A. A scaling normalization method for differential expression analysis of RNA-seq data. Genome Biol 11, R25 (2010).

52. Law, C.W., Chen, Y., Shi, W. & Smyth, G.K. voom: Precision weights unlock linear model analysis tools for RNA-seq read counts. Genome Biol 15, R29 (2014).

53. Ritchie, M.E. et al. limma powers differential expression analyses for RNA-sequencing and microarray studies. Nucleic Acids Res 43, e47 (2015).

54. Smyth, G.K. limma: Linear Models for Microarray Data, in Bioinformatics and Computational Biology Solutions Using R and Bioconductor. (eds. R. Gentleman, V.J. Carey, W. Huber, R.A. Irizarry & S. Dudoit) 397–420 (Springer New York, New York, NY; 2005).

55. Hotelling, H. Relations Between Two Sets of Variates, in Breakthroughs in Statistics: Methodology and Distribution. (eds. S. Kotz & N.L. Johnson) 162–190 (Springer New York, New York, NY; 1992).

56. Witten, D.M., Tibshirani, R. & Hastie, T. A penalized matrix decomposition, with applications to sparse principal components and canonical correlation analysis. Biostatistics 10, 515–534 (2009).

57. Tibshirani, R., Hastie, T., Narasimhan, B. & Chu, G. Diagnosis of multiple cancer types by shrunken centroids of gene expression. Proc Natl Acad Sci U S A 99, 6567–6572 (2002).

58. Wishart, D.S. et al. DrugBank 5.0: a major update to the DrugBank database for 2018. Nucleic Acids Res 46, D1074–d1082 (2018).

59. Colaprico, A. et al. TCGAbiolinks: an R/Bioconductor package for integrative analysis of TCGA data. Nucleic Acids Res 44, e71 (2016).

60. Robinson, M.D., McCarthy, D.J. & Smyth, G.K. edgeR: a Bioconductor package for differential expression analysis of digital gene expression data. Bioinformatics (Oxford, England) 26, 139–140 (2010).

61. Flamary, R., Courty, N., Rakotomamonjy, A. & Tuia, D. in NIPS 2014, Workshop on Optimal Transport and Machine Learning (Montréal, Canada; 2014).

62. Lin, H. & Zelterman, D. Modeling Survival Data: Extending the Cox Model. Technometrics 44, 85–86 (2002).

63. Kassambara, A., Kosinski, M. & Biecek, P. survminer: Drawing Survival Curves using ‘ggplot2’. R package version 0.3, 1 (2021).

64. Hu, H. et al. AnimalTFDB 3.0: a comprehensive resource for annotation and prediction of animal transcription factors. Nucleic Acids Res 47, D33–d38 (2019).

65. Orchard, S. et al. Protein interaction data curation: the International Molecular Exchange (IMEx) consortium. Nature methods 9, 345–350 (2012).

66. McQuin, C. et al. CellProfiler 3.0: Next-generation image processing for biology. PLoS biology 16, e2005970 (2018).

67. Berthold, M. et al. KNIME: The Konstanz Information Miner, in Data Analysis, Machine Learning and Applications; Studies in Classification, Data Analysis, and Knowledge Organization (Springer, 2007).

68. Kluyver, T. et al. in 20th International Conference on Electronic Publishing (01/01/16). (eds. F. Loizides & B. Scmidt) 87–90 (IOS Press.

69. Hunter, J.D. Matplotlib: A 2D Graphics Environment. Computing in Science & Engineering 9, 90–95 (2007).

70. Harris, C.R. et al. Array programming with NumPy. Nature 585, 357–362 (2020).

71. Virtanen, P. et al. SciPy 1.0: fundamental algorithms for scientific computing in Python. Nature methods 17, 261–272 (2020).

72. Waskom, M.L. seaborn: statistical data visualization. Journal of Open Source Software 6, 3021 (2021).

73. Walter, W., Sánchez-Cabo, F. & Ricote, M. GOplot: an R package for visually combining expression data with functional analysis. Bioinformatics (Oxford, England) 31, 2912–2914 (2015).

74. Yu, G., Wang, L.G., Han, Y. & He, Q.Y. clusterProfiler: an R package for comparing biological themes among gene clusters. Omics 16, 284–287 (2012).

