## Supplemental Figures 1-7 for "Targeting phenotypic plasticity prevents metastasis and the development of chemotherapy-resistant disease"

**Supplementary Information**

This document contains:

Supplementary Figure 1
Supplementary Figure 2
Supplementary Figure 3
Supplementary Figure 4
Supplementary Figure 5
Supplementary Figure 6
Supplementary Figure 7

#### SUPPLEMENTARY FIGURES AND LEGENDS

##### Supplementary Figure 1

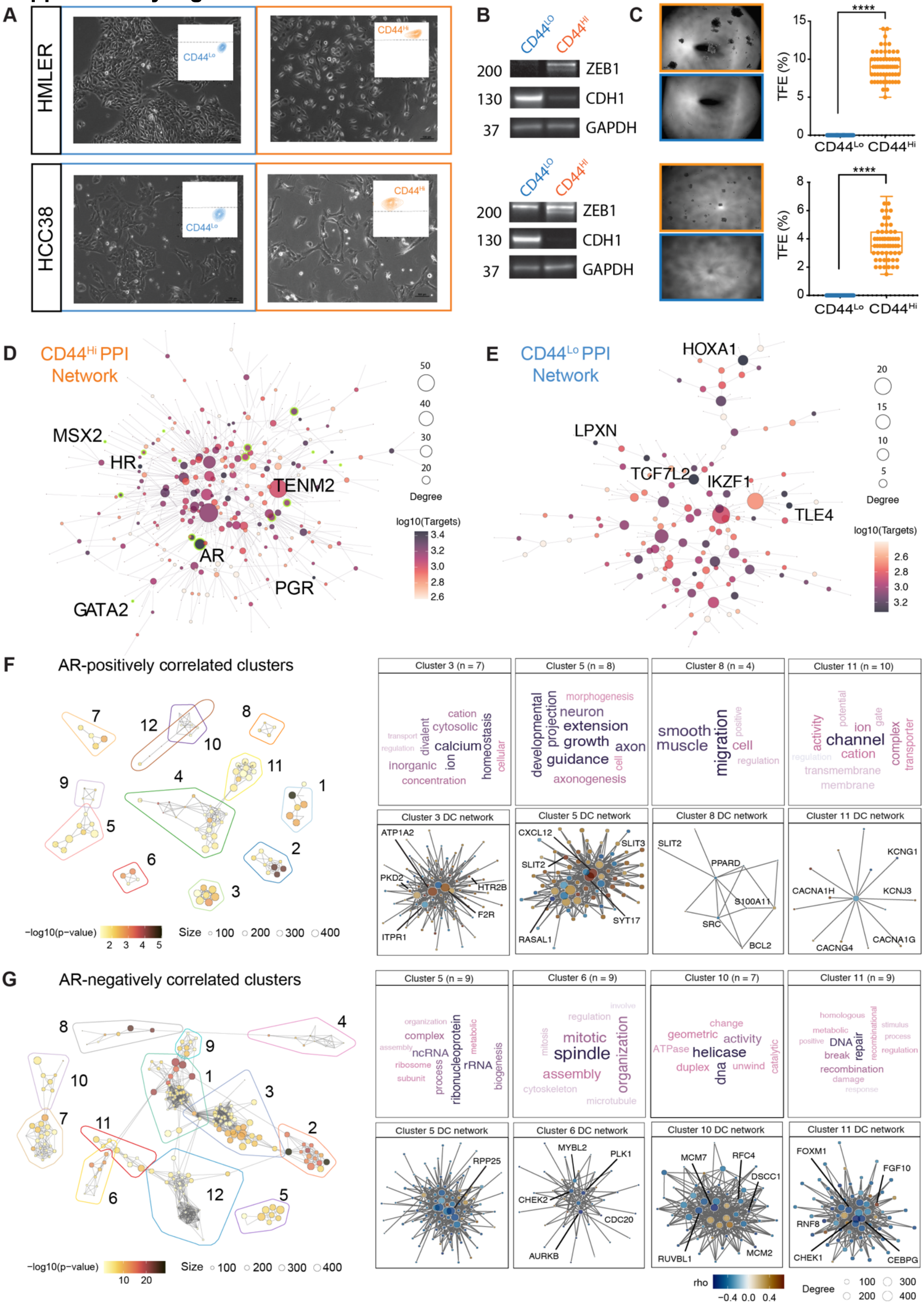

***Supplementary Figure 1:***

***A,*** *Photomicrographs of HMLER and HCC38 cells showing cell morphology of pure* $CD44^{Lo}$ *and* $CD44^{Hi}$ *cells. Insert – representative FACS plot showing CD44 (y-axis).* ***B,*** *Western blot showing expression levels of ZEB1, CDH1 (E-cadherin) and GAPDH in* $CD44^{Lo}$ *and* $CD44^{Hi}$ *cell populations.* ***C,*** *Photomicrographs (left) and quantification (right) of tumoursphere formation in matched* $CD44^{Lo}$ *and* $CD44^{Hi}$ *cells.* ***D,*** *A differential regulatory network of signalling pathways enriched in* $CD44^{Hi}$*-like TCGA breast cancer samples.* ***E,*** *A differential regulatory network of signalling pathways enriched in* $CD44^{Lo}$*-like TCGA breast cancer samples.* ***F,*** *Androgen receptor (AR) protein-protein cluster interactions positively correlated in TCGA* $CD44^{Hi}$*-like samples.* ***G,*** *AR protein-protein cluster interactions negatively correlated in TCGA* $CD44^{Hi}$*-like samples.*

### Supplementary Figure 2

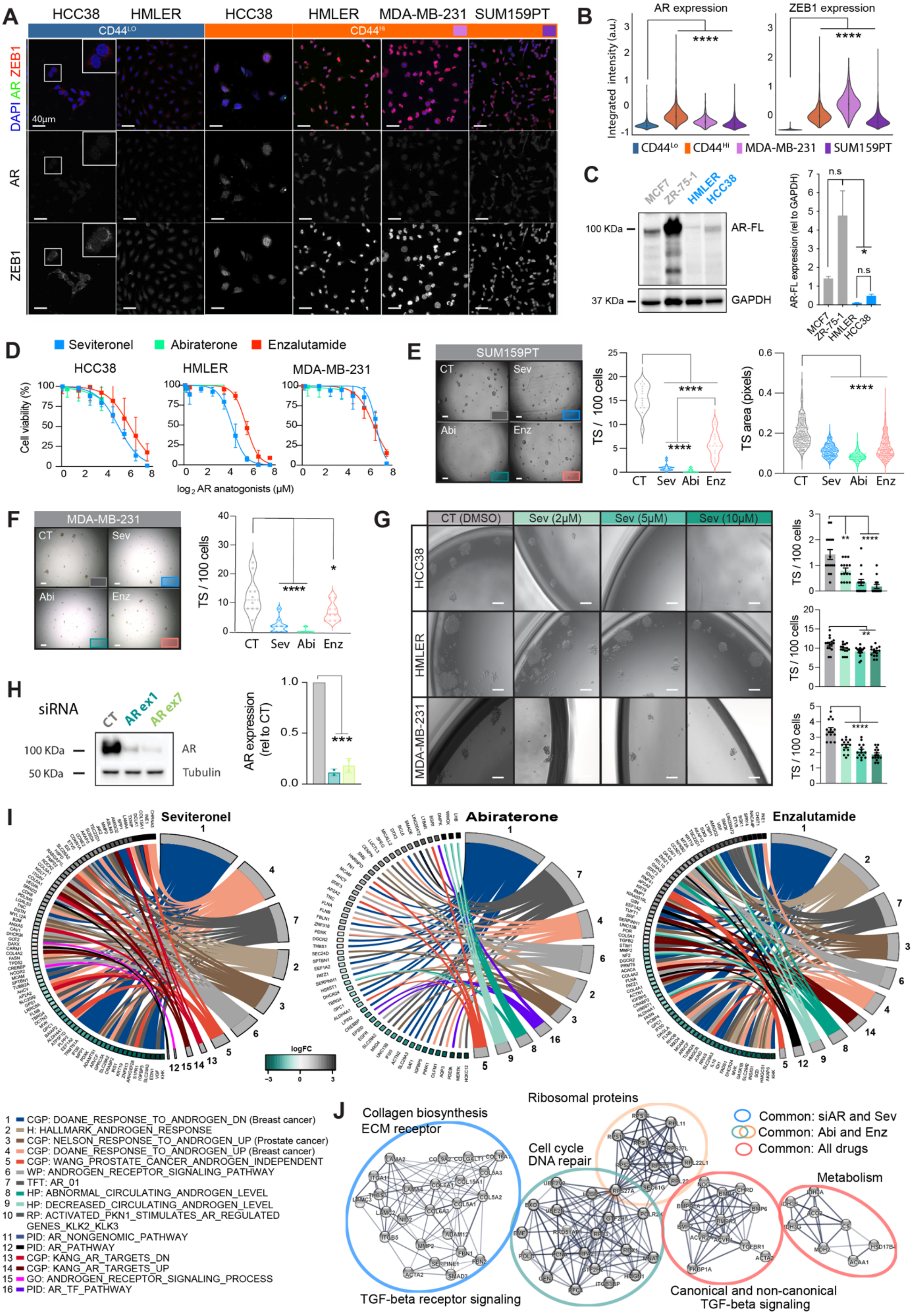

**Supplementary Figure 2:**

**A,** Immunofluorescence (IF) staining for AR (green), ZEB1 (green) and DAPI (blue) in various $CD44^{Lo}$ and $CD44^{Hi}$ cell subpopulations. **B,** Quantification of AR and ZEB1 levels from A. **C,** Dose response curves for seviteronel (blue), Abiraterone (green) and Enzalutamide (red) in $CD44^{Hi}$ subpopulation of the indicated cell lines. **D,** Western blot and quantification of AR protein in luminal and basal breast cancer cell lines. **E,** Photomicrographs (left) and quantification (right) of MDA-MB-231 tumoursphere-formation in response to Vehicle (DMSO), Seviteronel (Sev), Abiraterone (Abi) or Enzalutamide (Enz). **F,** Photomicrographs (left) and quantification (right) of tumoursphere-formation in response to decreasing dose of Seviteronel in various basal $CD44^{Hi}$ cell populations. **G,** Western blot (left) and quantification (right) or AR protein expression in Control (CT) or siAR (ARex1 and ARex7) 48h post transfection. **H.** AR targeted genes defined for each treatment condition matching other MSigDB AR signatures (continuation of Fig. 2I). AR targets selected from the top 1000 differentially expressed genes from the comparison between Sev, Abi or Enz versus vehicle (DMSO). **I.** String protein network analysis showing the similarities and differences in up- (UP) and down- (DN) regulated protein interactions from Reactome Pathways (Fig. 2K) enriched in siAR compared to siCT, and Sev, Abi or Enz compared to vehicle (DMSO).

#### Supplementary Figure 3

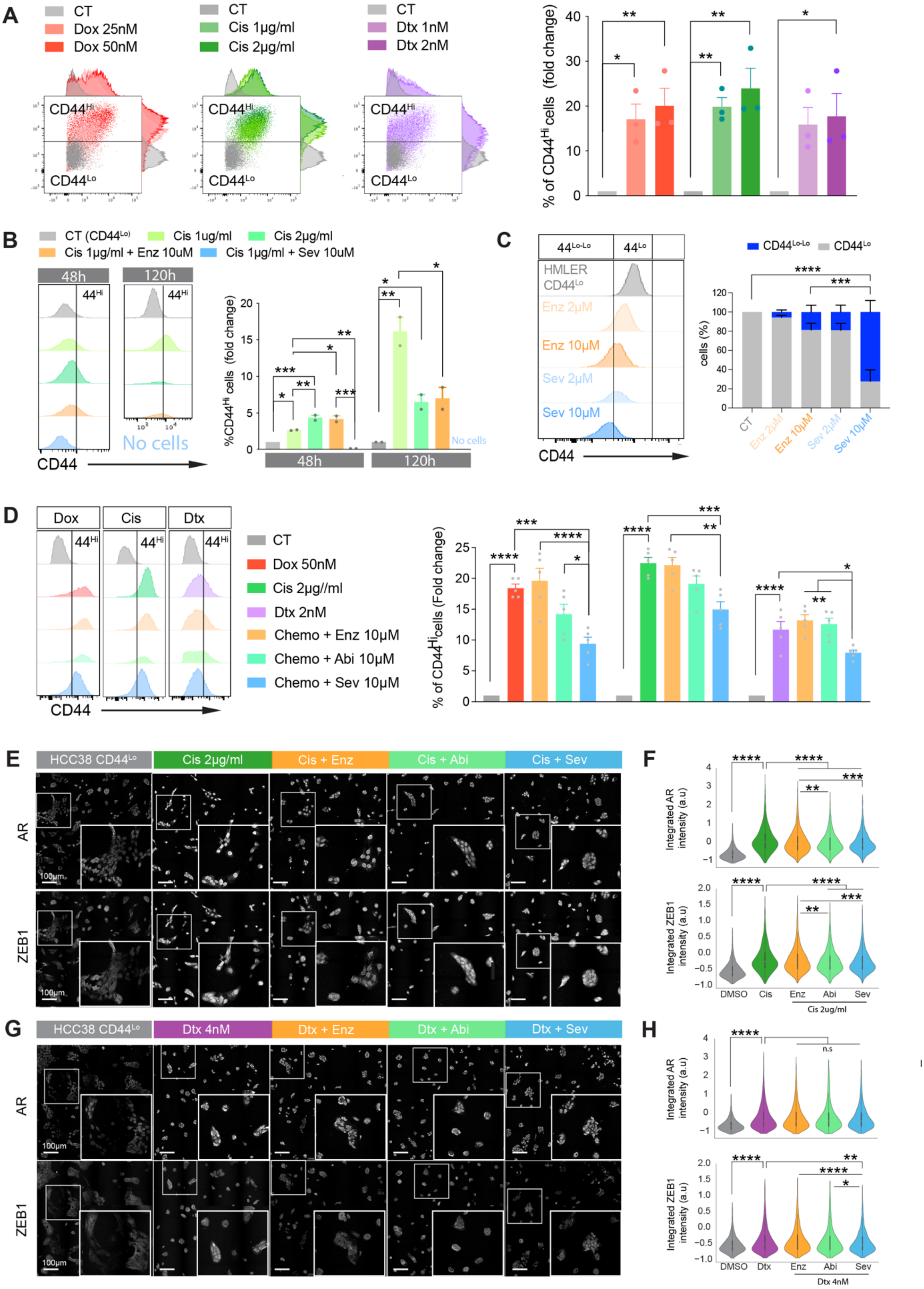

***Supplementary Figure 3:***

***A,*** *FACS analysis of dose-dependent induction of $CD44^{Lo}$ to $CD44^{Hi}$ cell state switching by Doxorubicin (Dox), Cisplatin (Cis) and Docetaxel (Dtx) in HCC38 cells 144h in treatment. Right bar plot - quantification of newly-generated $CD44^{Hi}$ cells.* ***B,*** *FACS analysis of $CD44^{Lo}$ to $CD44^{Hi}$ cell state switching in HMLER cells at 48h and 120h following treatment with CT, Cis, Cis + Enz or Cis + Sev (histograms, left) and quantification (bar plots, right).* ***C,*** *FACS analysis (Histograms, left) of CD44 status in HMLER $CD44^{Lo}$ cells treated with Enz or Sev alone, and, quantification of $CD44^{Lo\text{-}Lo}$ cells (bar plots, right).* ***D****, FACS analysis of $CD44^{Hi}$ status in HCCC38 cells for 144 hours following treatment with CT, Dox, Cis or Dtx, plus or minus Enz, Abi, Sev (histograms, left) and quantification (bar plots, right).* ***E-H,*** *Immunofluorescent images of AR and ZEB1 staining in HCC38 $CD44^{Lo}$ cells treated for 48h with chemotherapy (Cisplatin 2ug/ml or Docetaxel 4nM) with or without Sev, Abi or Enz (E, J) and quantification (F, H).*

#### Supplementary Figure 4

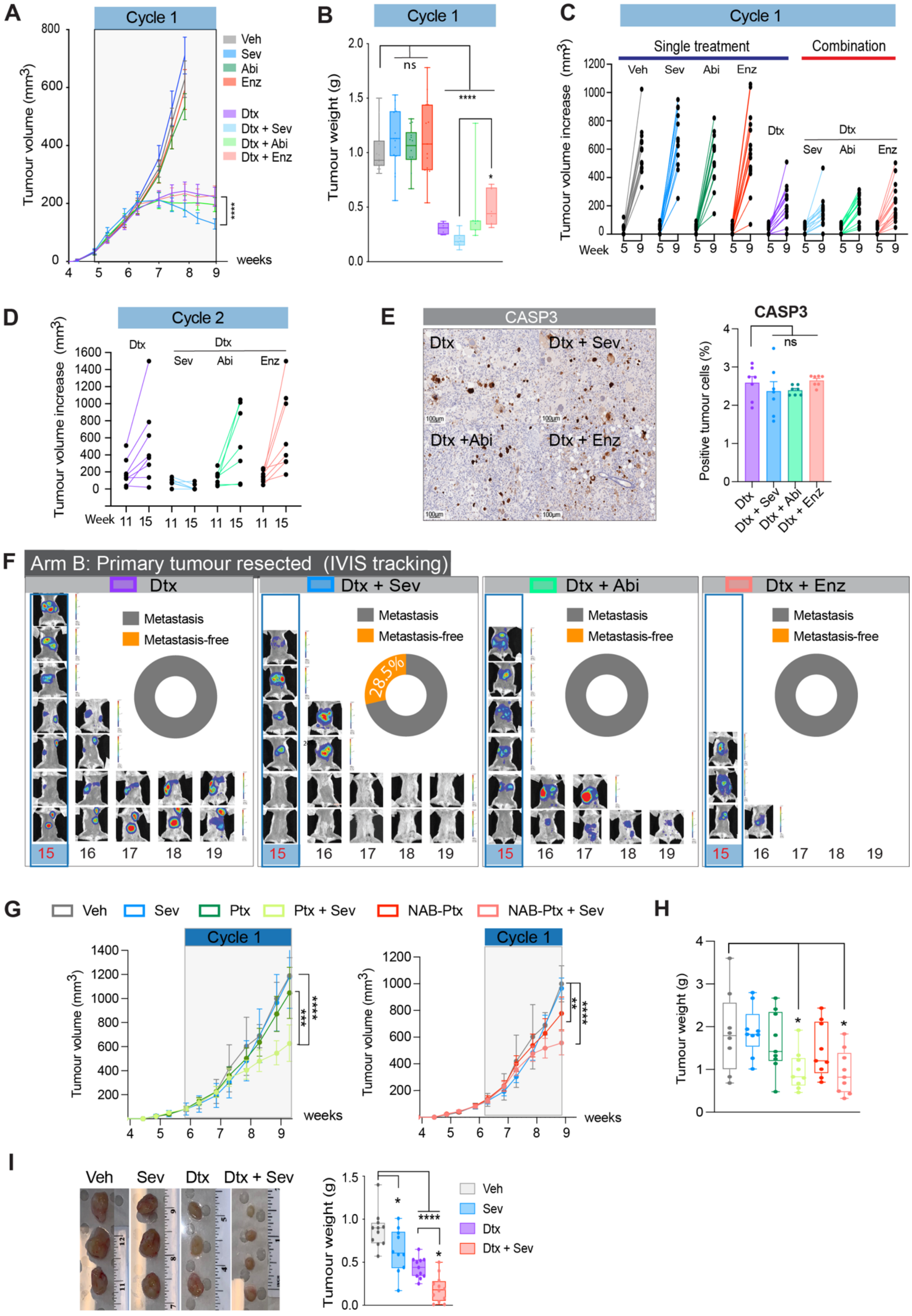

***Supplementary Figure 4:***

***A,** Growth kinetics of MDA-MB-231 cells treated with vehicle (DMSO 0.1% v/v, n = 15), Docetaxel (Dtx 20 mg/ml, n = 15), single-agent Sev, Abi, Enz (100 mg/kg/day for each AR antagonist, n = 15 animals per group), or, Dtx + Sev, Dtx + Abi or Dtx + Enz. **B,** Final tumour weights of animals harvested in Arm B at end of cycle 1. **C,** Comparison of growth evolution from week 5 to week 9 during Cycle 1 for all treatment groups in A. **D,** Comparison of growth kinetics from week 11 to week 15 during Cycle 2 for all combination treatment groups in A. **E,** Immunohistochemistry staining and quantification for cell death marker CASP3 in Dtx, Dtx + Sev, Dtx + Abi or Dtx + Enz treated tumours in Arm B harvested at end of cycle 1. **F,** IVIS imaging for luciferase to identify metastases in animals with primary tumour resected over weeks 15 to 19. Insert shows percentage of metastasis-free animals at the completion of the experiment. **G,** Growth kinetics of MDA-MB-231 tumours following treatment for 1 cycle with veh, sev, paclitaxel (Ptx), Ptx + Sev, albumin bound NAB-paclitaxel (NAB-Ptx) or NAB-Ptx + Sev. **H,** Final tumour weight from G. **I,** Left: Representative photographs of tumours treated with veh, sev (where first dose of Sev was 150 mg/kg), Dtx, or Dtx + Sev. Right: quantification of final tumour weights, (n = 12/group).*

**Supplementary Figure 5**

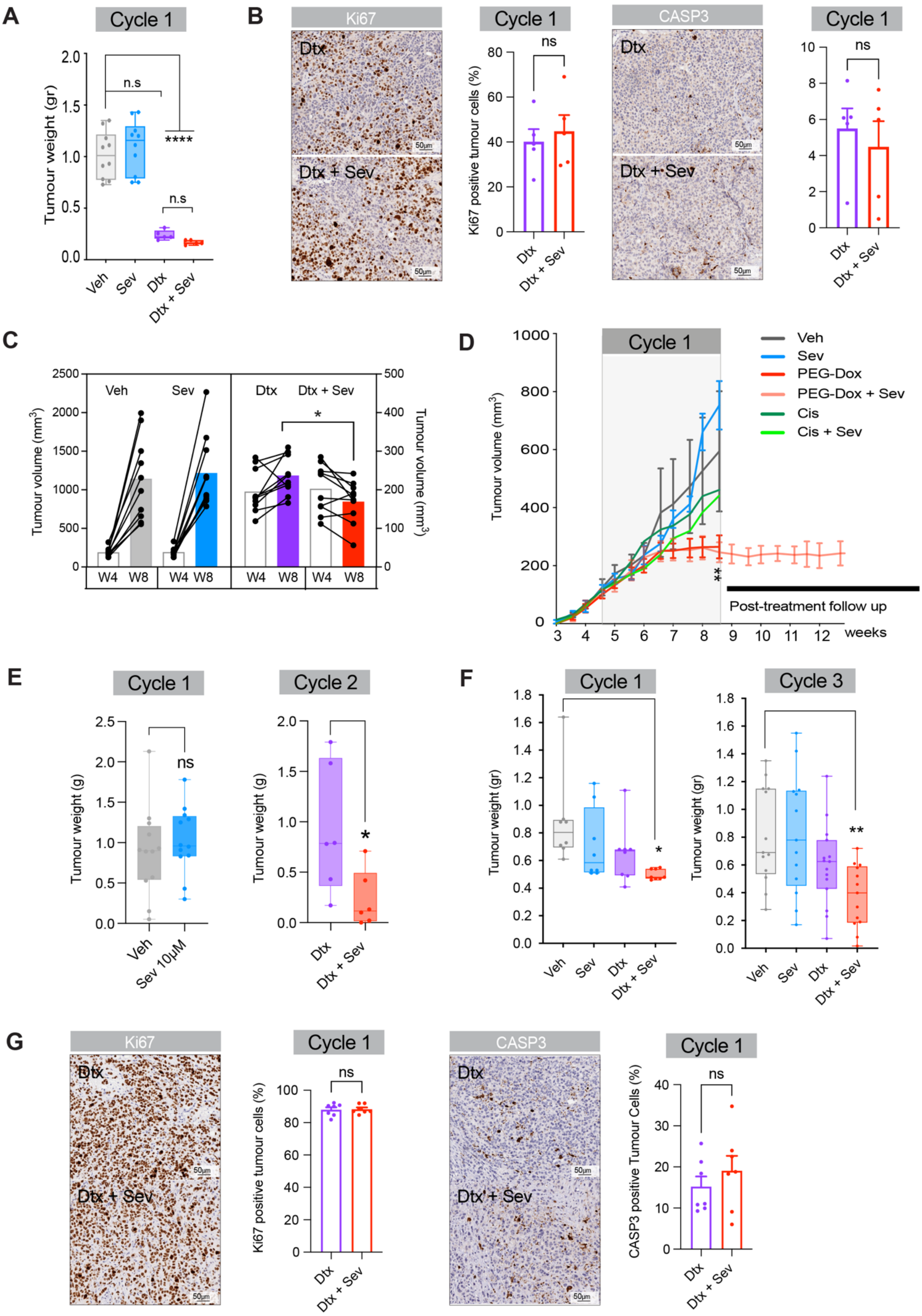

**Supplementary Figure 5:**

***A,*** *Final tumour weights of PDX Gar12-58 tumours harvested at the end of cycle 1 following treatment with vehicle (Ve, n = 10 mice), Seviteronel, (Sev; n = 10 mice), Docetaxel (Dtx, n = 6 mice), or Dtx + Sev (n = 6 mice).* ***B,*** *Immunohistochemistry on tumours treated with Dtx or Dtx + Sev with quantification (bar graph) stained with a marker for proliferation (Ki67 – LEFT, n = 5 tumours) or apoptosis (caspase 3 (CASP3) – RIGHT, n=5 tumours).* ***C,*** *Comparison of change in tumour volume from week 4 to week 8 in animals treated with vehicle (Veh), Dtx, Sev, or Dtx + Sev.* ***D,*** *PDX Gar12-58 tumour growth kinetics in mice treated for 1 cycle with Veh, Sev, pegylated-Doxorubicin (PEG-Dox), Cisplatin (Cis), PEG-Dox + Sev, or, Cis + Sev.* ***E,*** *Final tumour weights of Gar12-58 tumours harvested at the end of cycle 1 (Veh and Sev (n = 12 per mice)) and end of cycle 2 (Dtx and Dtx + Sev (n = 6 mice per group).* **F,** *Final tumour weights of PDX HCI-010 tumours harvested at the end of cycle 1 or cycle 3 following treatment with Veh, Sev, Dtx, or Dtx + Sev (n = 7 tumours per group and time point)*. **G,** *Immunohistochemistry for proliferation marker Ki67 on tumours treated with 1 cycle of DTX or DTX + Sev with quantification (bar graph). G, Immunohistochemistry for apoptosis marker caspase 3 (CASP3) (n = 7 tumours).*

#### Supplementary Figure 6

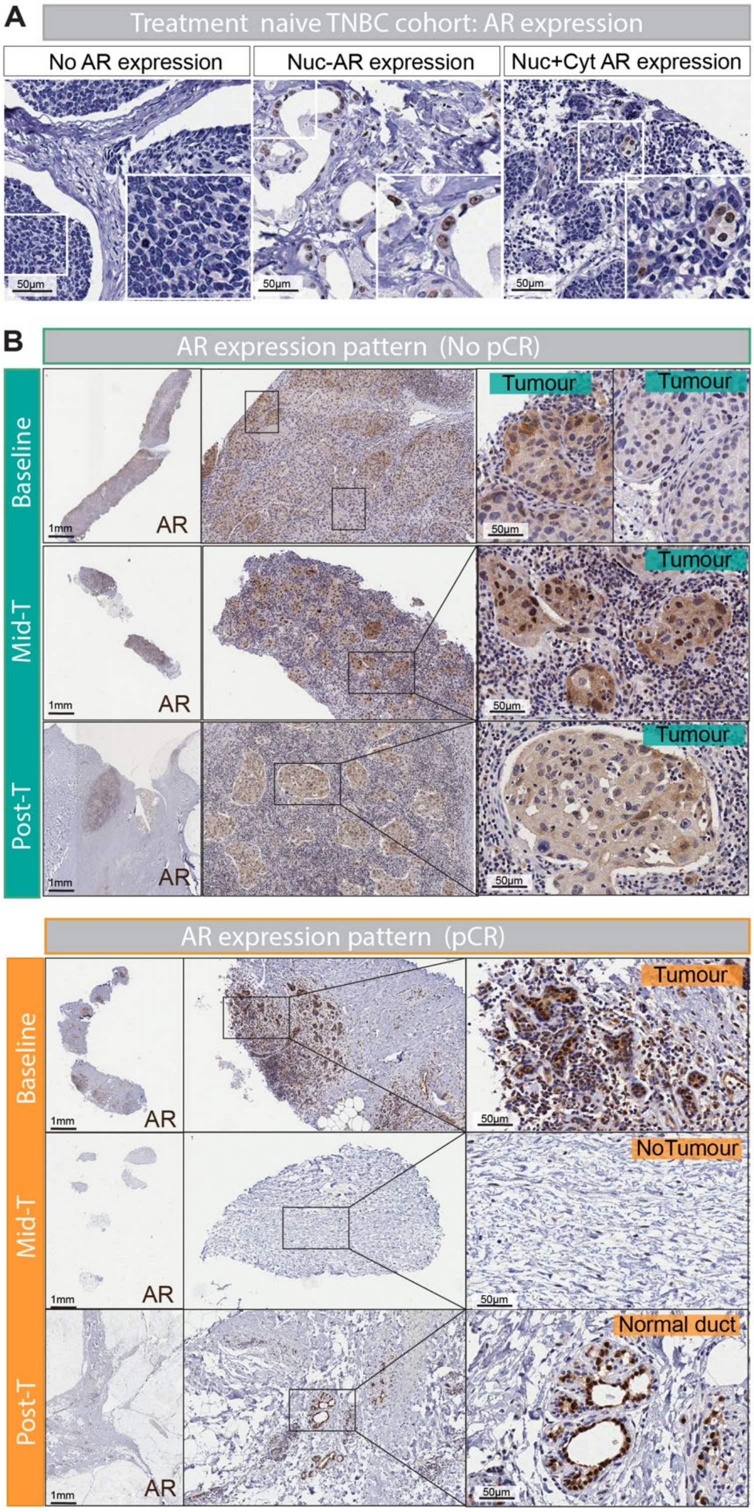


**C** SET UP TRIAL SAMPLE RECORD

| Patient | Baseline | Mid-T | Post-T | Outcome |
|---|---|---|---|---|
| 1 | 0 | 1 | 0 | no pCR |
| 2 | 1 | 1 | 1 | no pCR |
| 3 | 1 | 0 | 1 | no pCR |
| 4 | 1 | 0 | 0 | no pCR |
| 5 | 1 | 1 | 0 | no pCR |
| 6 | 1 | 0 | 1 | no pCR |
| 7 | 0 | 1 | 0 | no pCR |
| 8 | 1 | 1 | 1 | no pCR |
| 9 | 1 | 1 | 1 | no pCR |
| 10 | 1 | 0 | 0 | no pCR |
| 11 | 1 | 1 | 1 | no pCR |
| 12 | 1 | 1 | 1 | no pCR |
| 13 | 0 | 1 | 1 | no pCR |
| 14 | 0 | 1 | 1 | no pCR |
| 15 | 0 | 0 | 1 | no pCR |
| 16 | 1 | 0 | 1 | no pCR |
| 17 | 1 | 1 | 0 | no pCR |
| 18 | 0 | 1 | 0 | no pCR |
| 19 | 1 | 0 | 0 | no pCR |
| 20 | 0 | 1 | 1 | no pCR |
| 21 | 1 | 0 | 0 | pCR |
| 22 | 1 | 1 | 0 | pCR |
| 23 | 1 | 0 | 1 | pCR |
| 24 | 0 | 1 | 0 | pCR |
| 25 | 0 | 1 | 0 | pCR |
| 26 | 1 | 0 | 0 | pCR |
| 27 | 1 | 0 | 0 | pCR |
| 28 | 1 | 0 | 0 | pCR |
| 29 | 1 | 0 | 0 | pCR |
| 30 | 1 | 0 | 0 | pCR |
| 31 | 1 | 1 | 0 | pCR |

***Supplementary Figure 6:***

***A,*** *Immunohistochemistry images of androgen receptor (AR) staining showing negative, nuclear-only and nuclear plus cytoplasmic staining in cancer cells in treatment-naïve triple-negative breast cancer specimens.* ***B,*** *Representative immunohistochemistry images at low, medium and high magnification of AR staining (pre-treatment), during treatment (Mid-T) and post-treatment (Post-T) in patients from the SET-UP trial. Green cohort: images from a patient that did not obtain a pathological complete response (No pCR). Orange cohort: images from a patient that did obtain a pathological complete response (pCR). NB: AR positive staining in Post-T image is normal breast tissue.* ***C,*** *Individual patient tumour tissue availability and patient outcome. '0' = no tumour tissue available for analysis, '1' – tumour tissue available for analysis.*

#### Supplementary Figure 7

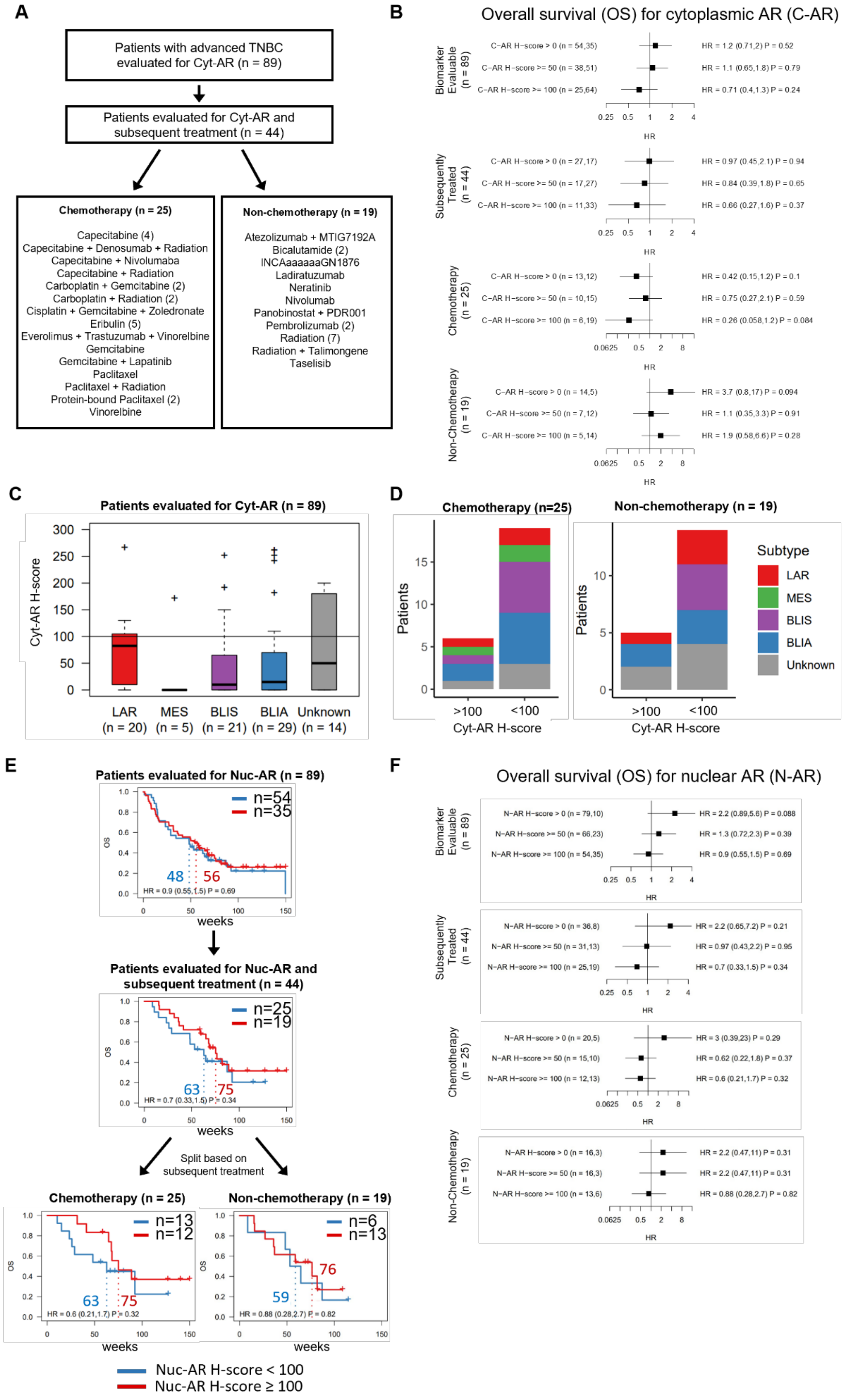

***Supplementary Figure 7:***

***A,*** *Flowchart showing chemotherapy and non-chemotherapy treatments for patient populations represented in Figure 7.* ***B,*** *Forest plots showing overall survival (OS) for cytoplasmic AR H-score cut-offs of >0, >=50 and >=100 in patient populations represented in Figure 7.* ***C,*** *Representation of Cyt-AR expression (H-score) according to TNBC molecular subtypes(1) of the 89 patient tumours in the Clarity-01 clinical trial (from Figure 7A).* ***D,*** *Representation of the TNBC molecular subtypes of tumours in the high and low Cyt-AR tumours (Clarity-01 trial) within the chemotherapy and non-chemotherapy subsets.* ***E,*** *Overall survival analysis of the Phase II Clarity-01 clinical trial. Kaplan-Meier plots for overall survival (OS) in patients treated with Seviteronel monotherapy split into low and high nuclear AR (Nuc-AR) expression based on H-score < 100 or ≥ 100. HR, hazard ratio with 95% confidence interval in brackets. Dotted lines indicate median survival times with 95% confidence interval in brackets.* ***F,*** *Forest plots showing overall survival (OS) for nuclear AR H-score cut-offs of >0, >=50 and >=100 in patient populations represented in Supp. Fig. 7C).*
